# Genetic Insight into the Paradoxical Relationship among Diabetes, Coronary Artery Disease, and Abdominal Aortic Aneurysm

**DOI:** 10.1101/2024.12.24.24319595

**Authors:** Shufen Zheng, Yonglin Wu, Aijie Li, Zhuoyi Wu, Zhen Liu, Huawei Wang, Xiaoyan Jia, Philip S. Tsao, Cuiping Pan

## Abstract

Type 2 diabetes (T2D) increases the risk of coronary artery disease (CAD) but decreases that for abdominal aortic aneurysm (AAA), forming an intriguing diabetes-atherosclerosis paradox. We investigate its genetic basis employing techniques such as genetic correlation, colocalization, gene annotation, functional enrichment, and pathway pairing on GWAS datasets. We discover a strong and positive correlation between T2D and CAD throughout the genome, with shared enrichment in immune signaling. The genetic correlation between CAD and AAA is weaker, with shared genetic components related to lipid metabolism. Conversely, T2D and AAA show the weakest genetic correlation, counter-balanced by two-thirds of genes and chromosomal segments with positive correlations and one-third with negative correlations. The positive correlations entail immune signaling, whereas the negative correlations are characteristic of beta-cell function and lipid metabolism. Our study suggests immune signaling contributes to the synergy between diabetes and atherosclerosis. By decoding the genetic interplay underlying these diseases, our findings provide a foundation for improving treatment strategies and advancing precision medicine.

## Introduction

Type 2 diabetes mellitus (T2D) is a recognized risk factor for cardiovascular diseases, with approximately one third of T2D patients world-wide presenting with cardiovascular comorbidity ^1^. Demonstrated in both clinical and animal studies, diabetes accelerates atherosclerosis through vasoconstriction, inflammation, and thrombosis ^2,3^. Consequently, one would expect a synergy for T2D and atherosclerotic diseases. However, epidemiological data indicate that T2D correlates positively with coronary artery disease (CAD) but negatively with abdominal aortic aneurysm (AAA) ^4^.

T2D is both a clinical and genetic risk factor for CAD ^5,6^. Mendelian Randomization (MR) studies support a causal role for T2D in the development of CAD ^7^. Investigations into shared genetic factors have identified mutations affecting vascular resistance and triglyceride metabolism ^8, 9^. Similarly, CAD is a well-established clinical and genetic risk factor for AAA. Prior studies, including our own, have indicated a bidirectional causal relationship between the two ^10,11^. Indeed, they share common risk factors including age, male gender, hypertension, dyslipidemia, and smoking ^12^, with a high co-prevalence observed in affected patients ^13^.

Despite the synergy between T2D and CAD, as well as CAD and AAA, a counter effect exists between T2D and AAA. Epidemiological meta-analyses indicate that T2D may protect against the incidence, growth, and prognosis of AAA ^14,15^. Multi-faceted mechanisms have been suggested for the protective effect of T2D against AAA, including endothelial dysfunction, chronic hyperglycemia, and insulin resistance ^16^. Essential genes and network proteins involved in these processes were identified ^17^. Meanwhile, several MR studies suggest that genetic predisposition to T2D confers no protection against AAA ^18,19^. Given the high heritability of T2D (72%) ^20^, AAA (77%) ^21^, and CAD (58-78%) ^22^, it remains to be determined whether genetic underpinnings dictate this inverse association.

In this study, we leverage recent large-scale genome-wide association studies (GWAS) to explore the genetic relationships of T2D, CAD, and AAA. We previously mapped the shared genetic architecture between AAA and 21 cardiometabolic traits, concluding that genetic predisposition to lipid metabolism and immune regulation largely defines the AAA pathogenesis ^11^. Here we apply it to study the genetic component of the diabetes-atherosclerosis paradox. This study unravels the genetic component of their intertwined relationships, offering insights for development of therapeutic strategies.

## Results

### Genetic predisposition reflects disease characteristics

We analyzed three recent large GWAS datasets for T2D ^23^, CAD ^24^, and AAA ^25^. Each study employed meta-analysis to interrogate more than 10 million SNVs in over 1 million individuals, resulting in 24,000, 26,000, and 6,700 genome-wide significant SNVs (*P* < 5 × 10^-8^) for T2D, CAD, and AAA, respectively (Supplementary Table 1). Power assessments suggest that all three datasets are sufficiently powered to detect SNVs with a minor allele frequency of 0.05 that influence disease risk by at least 10%. SNVs with lower allele frequency and larger effect sizes are also detectable (Supplementary Figure 1). Notably, several low-frequency SNVs were identified, with some exhibiting extreme odds ratios exceeding 10. For each disease, the inferred genes, pathways, tissues, and cell types well reflect disease characteristics (Supplementary Figure 2-4). For example, genetic signals of T2D are predominantly enriched in pancreatic cells, including beta, delta/gamma, and alpha cells, with additional strong signals linked to immune response. For the two atherosclerotic diseases, the genetic signals are notably enriched in the arteries and liver, as well as in pathways related to lipid metabolism. Thus, these GWAS datasets offer a robust foundation for investigating the genetic basis of the diabetes-atherosclerosis paradox.

### Shared genetic architecture among the three diseases

By integrating gene association results from GWAS (Methods), we derived 2,206 genes associated with T2D, 2,143 genes with CAD, and 860 genes with AAA. Consistent with epidemiological observations, T2D and CAD share the largest proportion of genes (Jaccard index (JI): 14.2%), followed by CAD-AAA (JI: 12.1%) and T2D-AAA (JI: 6.3%) (Supplementary Figure 5).

We previously developed a stringent framework to construct shared genetic architecture between diseases ^11^, which utilized cross-trait meta-analysis and gene-level association analysis to pinpoint shared SNVs and genes. Applying this framework, we assembled the shared genetic architecture among the three diseases (Figure 1A). Numerous genes, including *CDKN2B-AS1*, *CELSR2*, *CMIP*, *LPL*, *SLC22A3*, and *LOC646736*, are shared by all three disease pairs. This map also reflects disease-gene specificity, e.g., T2D predisposition gene *TCF7L2* ^26^ is identified for disease pairs involving T2D but not for CAD-AAA, and the atherosclerotic driver gene *ADAMTS7* ^27^ is identified for CAD-AAA but not for T2D disease pairs. Interestingly, *HMGCR*, *PCSK9*, and *BSN* ^28^ are identified for CAD-AAA only, whereas *APOE* is identified only for the T2D disease pairs. These findings suggest that lipid-related genes have distinct roles in the pathogenesis of the three diseases. Overall, T2D and AAA share much fewer SNVs and genes compared with other disease pairs.

**Figure 1.**
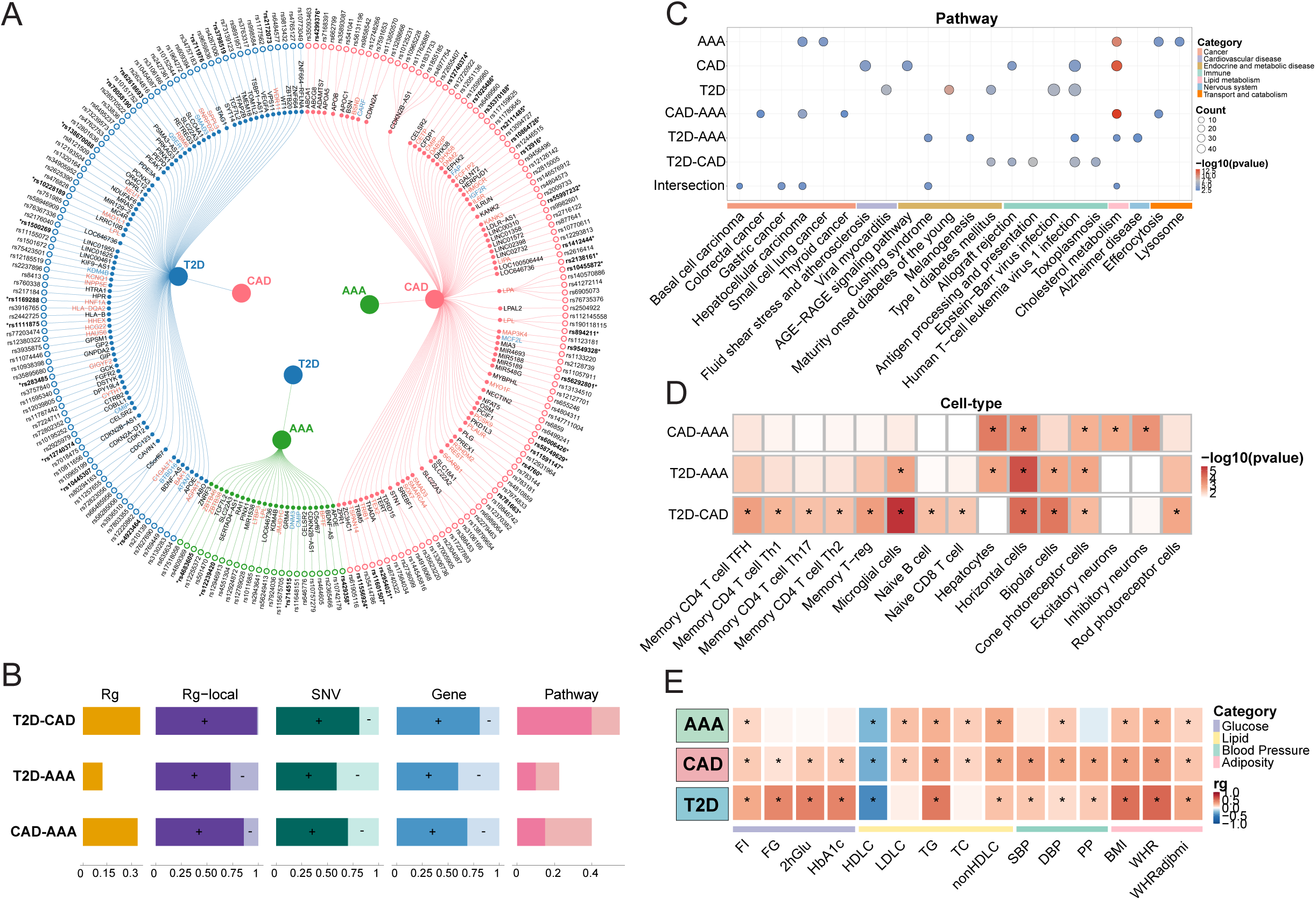
Assessment of genetic correlations among the three diseases. **(A)** SNVs and genes shared between diseases. Casual variants are marked by *. Genes names are color-coded: red indicates a positive association, blue indicates a negative association, and black marks unknown associations. **(B)** Summary of pairwise disease comparisons. Rg: genome-wide genetic correlation values. Rg-local: proportion of chromosomal segments with positive (dark color, “+”) or negative (light color, “-”) correlations between the diseases. SNV: proportion of shared GWAS SNVs with consistent (dark color, “+”) or inconsistent (light color, “-”) effects across diseases. Gene: proportion of shared GWAS genes with consistent (dark color, “+”) or inconsistent (light color, “-”) effects based on TWAS and SMR analysis. Pathway: pathway pairing scores, decomposed into tier-1 (dark color) and tier-2 (light color) pairing. **(C)** KEGG pathway enrichment for different sets of GWAS genes. The “intersection” set includes genes common to all three diseases. **(D)** Cell-type enrichment for different set of GWAS genes. Only results with P < 0.05 are displayed. *: P values passing the FDR-corrected threshold. **(E)** Genetic correlations between T2D/CAD/AAA and cardiometabolic traits. *: P values passing the Bonferroni-corrected threshold.

### T2D and CAD display a tight genetic correlation

We then quantified pair-wise genetic correlations for the three diseases (Figure 1B). CAD displays similar correlations with T2D (r_g_ = 0.35) and AAA (r_g_ = 0.34), while the correlation between T2D and AAA was markedly weaker (r_g_ = 0.12). Dividing the genome into 1 Mb segments, the local genetic correlations revealed a striking difference in association patterns. Of all correlated segments between T2D and CAD, 99% were positively correlated, with only 1% negatively correlated, indicating a strong concordance in association directions. Conversely, 27% of the segments between T2D and AAA were negatively correlated. This trend persisted when considering segments shared by all three diseases or those unique to specific disease pairs (Supplementary Table 2). In line with these observations, MR causal inference suggested bidirectional causality between T2D and CAD, and between CAD and AAA. No significant causality was inferred for T2D and AAA, although a marginal trend was observed in T2D against AAA (Supplementary Figure 6).

A SNV may denote increased risk for one disease but decreased risk for another. Grouping the shared SNVs by association direction, we derived a higher proportion of SNVs consistently associated with T2D and CAD (0.81), compared to CAD-AAA (0.70) and T2D-AAA (0.59) (Figure 1B). At the gene level, integration using transcriptome-wide association studies (TWAS) revealed a similar pattern: the proportion of genes with consistent association directions were higher for T2D-CAD (0.81) compared to T2D-AAA (0.60) and CAD-AAA (0.69). Therefore, not only a larger proportion of SNVs and genes are shared by T2D and CAD, but also, they are more aligned in direction of association.

The GWAS genes were enriched in specific biological pathways. Quantifying the pathway paring ^29^ derived a high matching score for T2D and CAD (score of 0.55) (Figure 1B). Importantly, this pairing occurred predominantly between the top enriched pathways for each disease (tier 1 pairing) (Supplementary Table 3). While CAD and AAA reached a moderate pathway paring (score of 0.40), most matches were in lower-rank pathways (tier 2 pairing). In contrast, T2D and AAA demonstrated a much weaker pathway connection (score of 0.22). Therefore, a tighter connection between T2D and CAD was found in biological pathways.

As expected, a strong enrichment in cholesterol metabolism was found for the atherosclerotic diseases (Figure 1C). Interestingly, immune response pathways were shared between T2D and CAD. Indeed, expression of their shared genes was enriched in T and B lymphocytes (Figure 1D). Meanwhile, the shared pathways and associated cell types for T2D-AAA and CAD-AAA were more diverse.

### T2D and CAD broadly correlate with metabolic traits

Correlation analysis indicated that CAD correlated with a broad catalogue of metabolic traits including adiposity, blood pressures, blood glucose, and blood lipids (Figure 1E). Consistent with their clinical characteristics, T2D showed strong correlations with glucose and adiposity traits, while AAA was primarily correlated with adiposity and blood lipids. These findings are consistent with causal inferences drawn from MR analyses (Supplementary Figure 7). Overall, the metabolic correlation profiles highlight that T2D and CAD share a more extensive genetic predisposition to metabolic regulation compared to other disease pairs.

From the above analyses we conclude that among the three diseases, T2D and CAD exhibit the strongest and most extensive genetic correlation, with the shared genes enriched for immune signals. The correlation between CAD and AAA is relatively weaker, characterized by sharing in lipid-related genes and pathways. T2D only displays a weak correlation with AAA. Taken together, genetic characteristics of the diseases align closely with observed patterns of disease co-occurrence.

### Gene sets that explain the negative correlation between T2D and AAA

A protective trend of T2D against AAA was observed (Supplementary Figure 6), warranting further investigation. We reanalyzed the relationship using different sets of genetic instruments derived from curated T2D SNVs ^30^ and two independent T2D GWAS ^23,31^, applying various MR models. These analyses reported odds ratios of 0.5-0.9 for T2D on AAA; a negative correlation was not observed when the causal direction was reversed (Figure 2A), suggesting T2D indeed protects against AAA.

**Figure 2.**
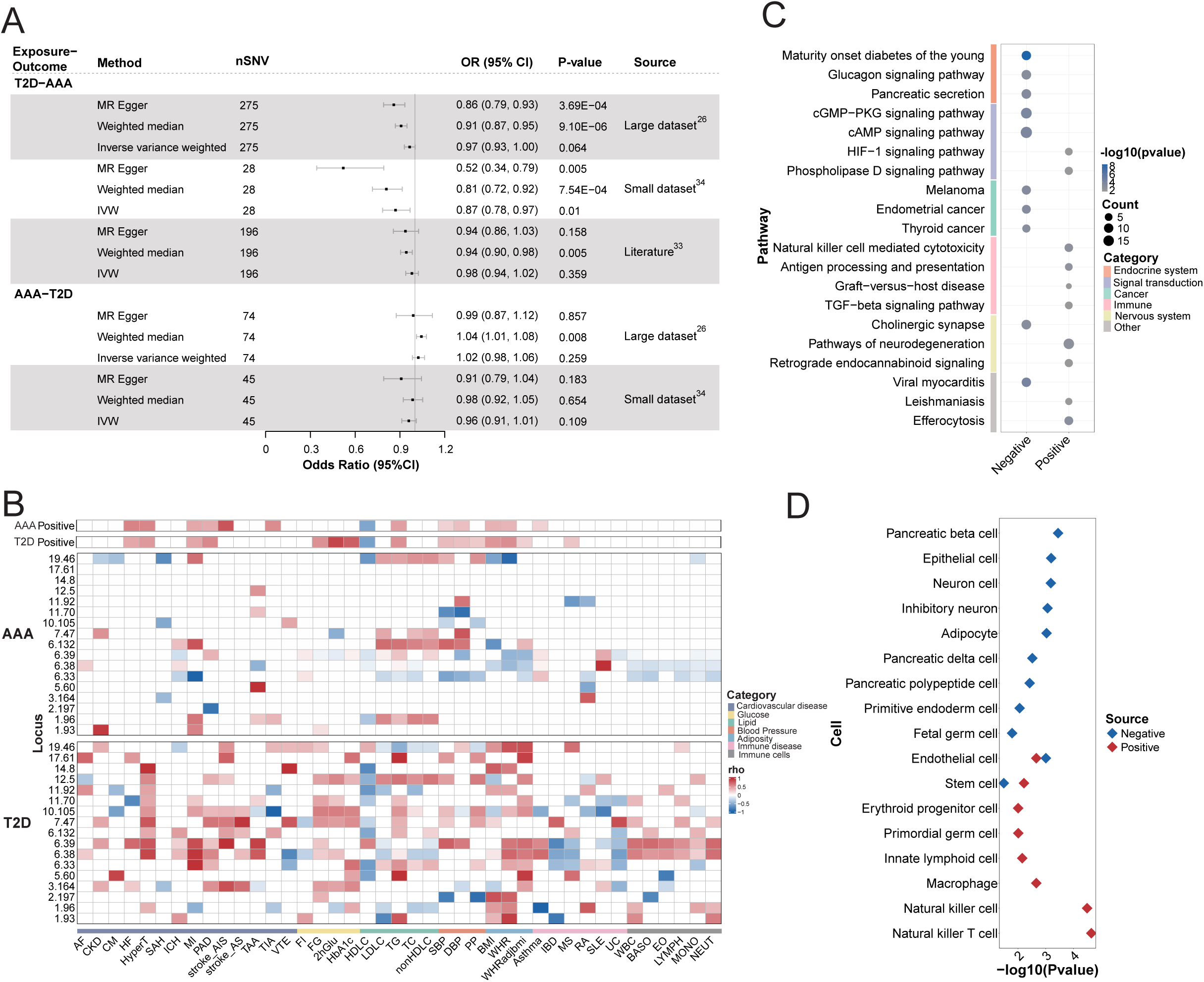
Explaining the inverse association between T2D and AAA. **(A)** Bidirectional MR analysis of T2D and AAA based on three sets of genetic instruments and three MR models. **(B)** Local genetic correlation profiles for T2D or AAA, with 29 cardiometabolic and 12 immune phenotypes. 46 positive correlation regions are combined to display an overall profile for AAA (“AAA Positive”) or T2D (“T2D Positive”), alongside 17 negative correlation segments, each presented individually. **(C)** KEGG pathway enrichment for gene sets exhibiting either positive or negative correlations between T2D and AAA. **(D)** Cell type enrichment for gene sets exhibiting positive or negative correlations between T2D and AAA. “Negative” refers to the combined set of negatively associated genes identified from three sources: annotations of MR instrumental variables, the 17 negatively correlated genomic regions, and TWAS/SMR analyses. “Positive” refers to the combined set of positively associated genes identified from two sources: the 46 positively correlated genomic regions and TWAS/SMR analyses.

To uncover what contributes to this negative correlation, we examined GWAS genes with opposite associations with the two diseases. Among the 182 genes shared by T2D and AAA, 173 genes were assigned association directions using TWAS, summary-data-based Mendelian randomization (SMR) and gene burden tests (Methods) (Supplementary Table 4). These genes were categorized into four groups based on their association directions. Notably, 38% exhibited opposite associations, i.e., AAA(-):T2D(+) and AAA(+):T2D(-), and demonstrated differential expression in affected tissues, i.e., the aorta in AAA patients (GSE57691) ^32^ and the pancreas in T2D patients (GSE38642) ^33^ (Supplementary Figure 8). In contrast, no expression differences were observed in the non-lesion tissue, such as the liver in T2D patients (GSE23343)^34^. These results suggest that the oppositely associated genes have direct functional relevance to the diseases.

### Chromosomal regions that underlie the negative correlation between T2D and AAA

We further searched for chromosomal regions that explain the inverse relationship. From local genetic correlation analysis, we identified 17 chromosomal segments with negative correlations and 46 segments with positive correlations between T2D and AAA. When examining disease correlations in these segments, including 29 cardiometabolic and 12 immunological phenotypes, we observed that T2D and AAA shared similar correlation profiles in the positive correlation segments. However, the profiles in the negative correlation segments were starkly different in that T2D correlated with many more phenotypes than AAA (Figure 2B).

We characterized the negative correlation segments by GWAS genes (Supplementary Figure 9). Locus 1.96 (chromosome 1, the 96^th^ segment) and locus 19.46 are enriched for lipid genes (e.g., *SORT1*, *PSRC1*, *CELSR*, *APOE, APOC1*, and *TOMM40*) and locus 7.47 is enriched for glucose and insulin signaling genes (e.g., *GCK* and *KCNJ3*). Furthermore, three segments, i.e., locus 6.33, locus 6.38, and locus 6.39, are dominated by HLA genes (Supplementary Figure 10). Conditioning on blood lipids or adiposity traits in these HLA segments led to a substantial reduction of genetic correlation between T2D and AAA (partial correlation analysis), implying lipid and adiposity drive the correlation. This was further confirmed by modeling AAA using T2D and lipid/adiposity traits as predictors (AAA ∼ T2D + blood lipids/adiposity) (multivariate regression) (Supplementary Table 5). As such, HLA immunity could impact lipid/adiposity traits, which in turn mediate the negative correlation between T2D and AAA. After annotating all genes in the negative correlation segments (Supplementary Table 6), we identified additional disease genes, including immune genes *IL18BP* and *TANK* in locus 11.70, and *TCF7L2* in locus 10.105.

### Pathways that explain the negative correlation between T2D and AAA

We then pooled all relevant genes to examine pathways and cell types. Genes for explaining the negative correlation between T2D and AAA include those derived from MR genetic instruments (n = 359), TWAS/SMR opposite associations (n = 43), and the negatively correlated segments (n = 362). They were enriched for endocrine functions (glucagon signaling and pancreatic secretion), cardiac contractility (cAMP and cGMP-PKG signaling), and growth hormone synthesis, secretion and action (Figure 2C, Supplementary Figure 11). Their expressions were enriched in various pancreatic cells, neurons, adipocytes, and endothelial cells (Figure 2D). In contrast, genes for explaining the positive correlation were enriched for immune response, with enriched expressions in natural killer cells, macrophages, and other immune cell types. Notably, the positive correlation signal of T2D and AAA largely resembles that of T2D and CAD, with an enrichment in immune response.

### Correlation of T2D mechanistic clusters with AAA and CAD

Eight mechanistic clusters of T2D have been reported, reflecting distinct disease etiologies ^35^. We interrogated their correlation profiles. For each T2D cluster, we identified the causal SNVs shared by CAD or AAA via colocalization. We note that the T2D SNVs shared with CAD were evenly distributed shared across the mechanistic clusters, whereas those shared with AAA were enriched in the lipodystrophy cluster and two beta-cell-dysfunction clusters (beta cell +PI cluster and beta cell -PI cluster, representing beta cell dysfunction with a positive or negative association with proinsulin) (Figure 3A). Strikingly, when we further grouped the shared SNVs by association directions, the two beta-cell-dysfunction clusters contain the largest proportion of SNVs that were oppositely associated with T2D and AAA (Figure 3B). This was not observed for the T2D-CAD disease pair. Furthermore, MR causal inference indicated the two beta-cell-dysfunction clusters of T2D were protective against AAA but harmful to CAD (Figure 3C). Taken together, our results from the gene, segment, and mechanistic cluster analyses suggest that the inverse relationship between T2D and AAA is driven by genetic predisposition to beta-cell functions and lipid metabolism, with the beta-cell functions being most influential.

**Figure 3.**
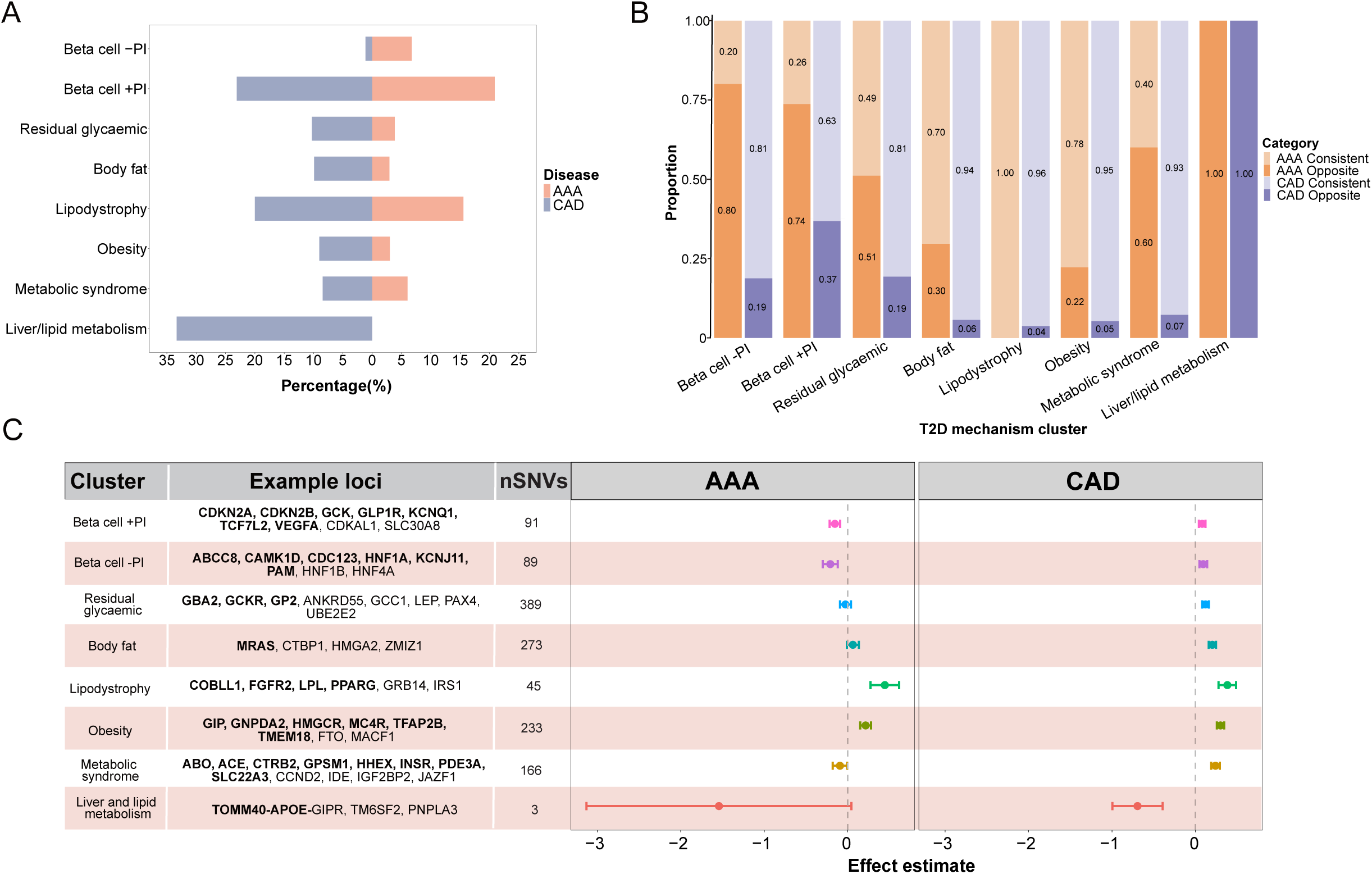
Beta-cell dysfunction underlies the inverse association between T2D and AAA. **(A)** Percentages of causal SNVs in each T2D mechanistic cluster that are shared with AAA (orange) or CAD (purple). PI: proinsulin. **(B)** Relative proportions of the shared SNVs in each T2D mechanistic cluster, categorized by their direction of associations, e.g., “AAA consistent” denotes the SNVs with aligned association directions with both T2D and AAA; “AAA opposite” denotes SNVs with opposite association directions with T2D and with AAA. **(C)** Causal effect of T2D mechanistic clusters on AAA or CAD. Clusters are displayed in “Cluster”. Example genes are provided in “Example Loci”. Total numbers of T2D GWAS SNVs are denoted in nSNVs. Effects of each T2D mechanistic cluster (exposure) on AAA or CAD (outcome) are presented in the forest plots.

### Genes and SNVs of opposite risks for T2D and AAA

For genes inferred to have opposite associations with T2D and AAA (Figure 4A), many indeed harbor SNVs with negative associations for one disease and positive associations for the other. Example GWAS association effects for several such genes are displayed (Figure 4B), including *CELSR2*, *APOE*, and *TOMM40* (lipid metabolism), *SMAD3* and *NUCB2* (TGF-β signaling, immunity), *HMGCR* and *KCNJ11* (drug targets), and *TCF7L2* (Wnt signaling, development). Their expressions were enriched in cardiometabolic axis, e.g., adipose tissue, skeletal muscle, liver, pancreas, cardiomyocyte, endothelial cell and hepatocytes (Supplementary Figure 12). Further colocalization analysis identified GWAS SNVs that correlated with gene expression (expression quantitative trait loci, eQTL), which are likely causal SNVs. As such, rs12740374 for *CESLR2*, rs34872471 for *TCF7L2*, and rs6453133 for *HMGCR* were identified (Figure 4C, Supplementary Table 7).

**Figure 4.**
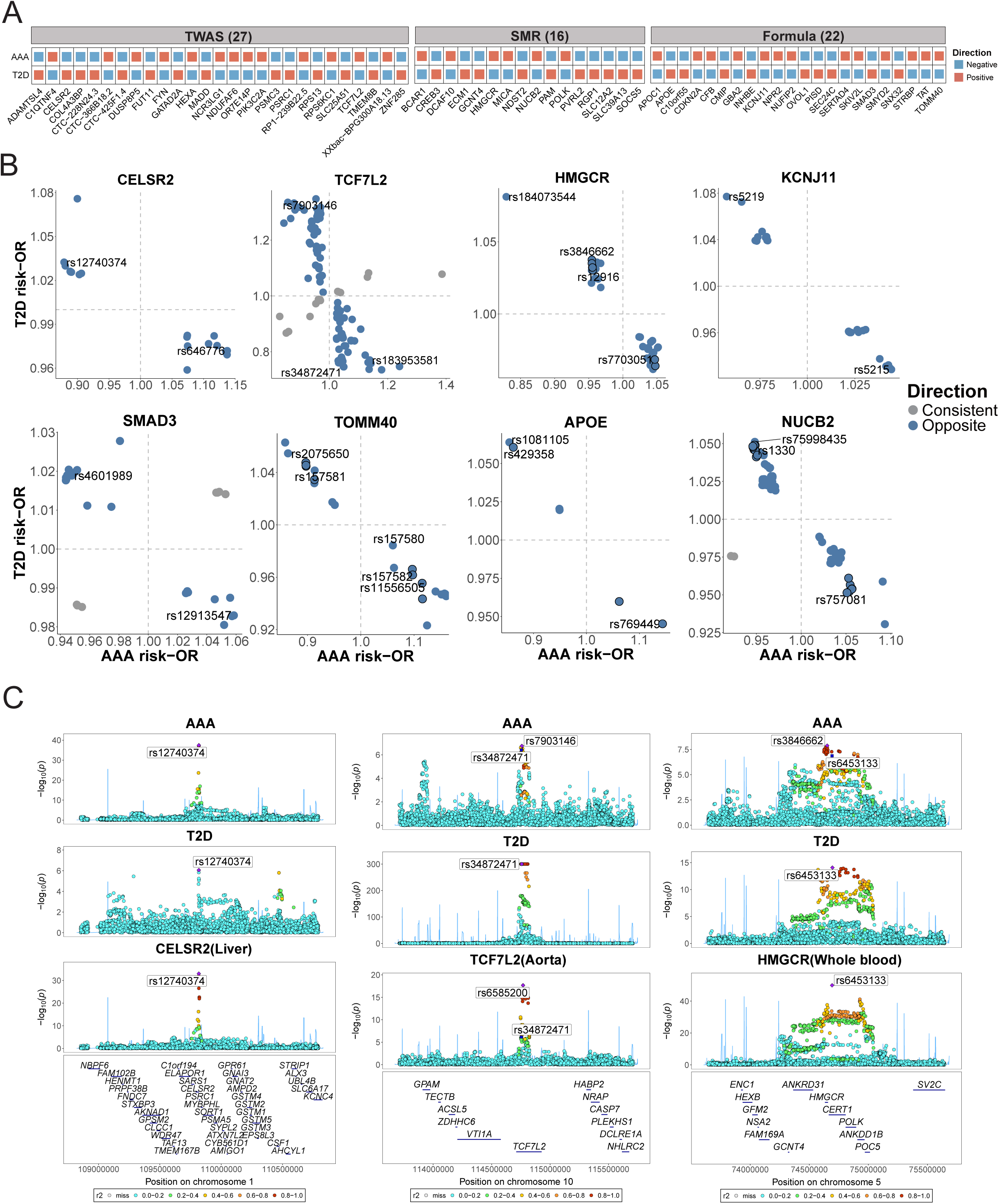
Pleiotropic effects of genes and SNVs on T2D and AAA. **(A)** Genes with opposite effects on T2D and AAA, derived from TWAS, SMR, or gene burden tests. **(B)** Odd ratios of SNVs shared between T2D and AAA (P < 0.05) on eight genes. SNVs with opposite associations with T2D and AAA are shown in blue, while those with consistent associations are shown in grey. **(C)** LocusZoom plots showing variants associated with AAA, T2D and gene expression. The labeled SNVs (with rsID) represent GWAS signals for T2D and AAA (first two rows) that are either tissue eQTLs or colocalize with tissue eQTLs (thrid row).

*CELSR2* is located in locus 1.96, a lipid-rich segment with a negative correlation between T2D and AAA. TWAS inferred that the increased *CELSR2* expression was associated positively with T2D but negatively with AAA, i.e., T2D(+):AAA(-). The T allele of rs12740374, a causal SNV for *CELSR2*, was associated with T2D(+) and AAA(-), potentially due to its effect on upregulating *CELSR2* expression in the liver (Supplementary Table 8). Phenome-wide association studies (PheWAS) linked rs12740374 to hyperlipidemia, hypercholesterolemia, and various cardiovascular diseases (Supplementary Figure 13A). Furthermore, rs12740374 is known to create a binding site for the transcription factor C/EBP and alter hepatic expression of *SORT1* ^36^. Therefore, this establishes a strong association with the CELSR2-PSRC1-SORT1 axis on chromosome 1p13.3, which is a critical pathway in lipid metabolism, particularly for low-density lipoprotein cholesterol (LDL-C). These results reinforce the role of lipid metabolism in mediating the negative correlation between the two diseases.

In contrast, *TCF7L2* in the locus 10.105 exhibited a T2D(-):AAA(+) association in the aorta. These suggest that a decreased *TCF7L2* expression in the aorta was associated with elevated risk of T2D and reduced risk of AAA. The C allele of rs34872471, a casual variant for *TCF7L2*, colocalized with an aortic eQTL which was associated with decreased *TCF7L2* expression. Conversely, the same allele displayed T2D(+):AAA(-):*TCF7L2* expression(-) (Supplementary Table 8). PheWAS linked this variant to diabetes and its complications (Supplementary Figure 13B). *TCF7L2* is known to encode for a transcriptional effector of the Wnt/β-catenin signaling pathway, which plays an important role in T2D pathogenesis ^37^. Although elevated expression of *TCF7L2* in pancreas has been associated with reduced risk of T2D ^38^, its higher expression in the aorta has been linked to increased risk of thoracic aortic aneurysm (TAA) ^39^. Our result in AAA aligns with that in TAA, indicating a consistent role of TCF7L2 in aortic aneurysms. Altogether, TCF7L2 is strongly suggested to play an opposing role in T2D and aortic aneurysms.

Lastly, the causal SNV rs6453133 for *HMGCR* is associated with T2D(-), AAA(+), and *HMGCR* expression(+). *HMGCR* encodes for 3-hydroxy-3-methylglutaryl-coA reductase, a rate-limiting enzyme for cholesterol synthesis and the target of lipid-lowering statins, and has been reported in numerous studies as a therapeutic target for treating aortic aneurysms ^40^. *HMGCR* was inferred in our SMR analysis as T2D(-):AAA(+) (Figure 4A), suggesting that increased expression of *HMGCR* was associated with reduced risk of T2D and increased risk of AAA. Therefore, inhibition of HMGCR could reduce the risk of AAA but potentially elevate the risk of T2D. In line with this, epidemiological observations have reported that intake of statins is associated with elevated risk of T2D ^41^. These results suggest that genetic studies could provide valuable insights into treatment effects.

## Discussion

In this study, we conducted a thorough interrogation of genetic relationships among T2D, CAD and AAA, confirming a genetic basis underlying the epidemiological observations of tight connections for T2D-CAD and CAD-AAA. At the molecular level, T2D and CAD shared many GWAS genes, display a significant overlap in their top-enriched pathways, and base their connection largely on immune response. CAD and AAA demonstrate strong enrichment in cholesterol metabolism, with shared gene expression predominately localized to hepatocytes. In contrast, the genetic relationship between T2D and AAA was notably weaker. Among their shared genetic components, 38% of genes and 27% chromosomal segments displayed negative correlations. Further investigation revealed that genes and regions with consistent direction of association with T2D and AAA were enriched for immune response, whereas those with opposite associations were primarily linked to beta-cell functions and lipid metabolism. In summary, our findings uncovered the genetic basis for the diabetes-atherosclerosis paradox (Supplementary Table 9).

Our study highlights the synergistic role of immune signaling in T2D and atherosclerosis. Previous reports have shown that hyperglycemia and dyslipidemia are linked to elevated levels of inflammatory cytokines, with T-cells playing a critical role in the development of diabetes and its complications ^42^. This aligns with our finding that shared genes between T2D and CAD show enriched expression in T-cells. Furthermore, diabetic patients with AAA display heightened inflammatory infiltration ^43^, and those with both diabetes and CAD exhibit increased local inflammation ^44,45^. Supporting these observations, a meta-analysis revealed that anti-inflammatory therapies in patients with comorbid T2D and CAD were associated with a reduced risk of major cardiovascular events ^46^. These findings suggest that targeting inflammatory pathways may hold therapeutic potential for managing T2D-related complications ^47,48^.

We also observed that genes exhibiting negative correlations between T2D and AAA were enriched in endocrine function, cardiac contractility, pancreatic cells, and endothelial cells. Among the eight T2D mechanistic clusters, the two clusters involving beta-cell dysfunction and associated with proinsulin levels (beta cell +PI cluster and beta cell -PI cluster) demonstrated a causal protective effect against AAA. In contrast, the residual glycemic cluster, which is not associated with proinsulin or decreased fasting insulin ^35^, showed no causal effect on AAA. These findings suggest that the genetic regulation of insulin dynamics, rather than hyperglycemia, plays a pivotal role in this protective effect. Notably, hyperglycemia may play an important role beyond genetic regulation. Indeed, multifaceted mechanisms attributed to hyperglycemia have been proposed for explaining the T2D protection against AAA, including glycation and cross-linking on extracellular matrix (ECM) proteins, reduced apoptosis of vascular smooth muscle cells (VSMCs), diminished intraluminal thrombus formation, decreased neovascularization, and attenuated inflammation^49^. Further research is warranted to delineate the distinct roles of insulin and glucose in mediating this protective effect.

In this study, lipid metabolism was identified as a shared genetic factor underlying CAD and AAA, as evidenced by their shared lipid-related genes, cell types, and signaling pathways. Consistent genetic correlations and causal effects between lipid traits and both CAD and AAA further support this finding. A previous study that interrogated genetic association network suggested that lipid metabolism represents a common biological pathway for eight vascular diseases, including CAD and AAA ^50^. Our results further emphasize the potential of lipid-lowering therapies as effective strategies for treating both CAD and AAA^51,52^.

Genetic features in tissue and cell type specificity provide further insights for targeted drug development in precision medicine. For both CAD and AAA, their genetic signals were enriched in arteries, liver, hepatocytes and endothelial cells. The positive genetic correlation signals between CAD and T2D, as well as T2D and AAA, were primarily enriched in immune cells, whereas the negative correlation signals between T2D and AAA were strongly associated with pancreatic beta cells. These findings underline the importance of leveraging disease-relevant tissues and cell types in drug development and efficacy prediction ^53^.

We recognize several limitations in our study. First, most of the input GWAS statistics were derived in European ancestry, which may limit the generalizability of our findings and provide only a partial view of the association landscape for the diseases. Second, while we inferred the direction of association between genes and diseases, functional studies are essential for validation and obtaining mechanistic insights, particularly regarding the inverse relationship between T2D and AAA. Third, beyond genetic factors, the regulatory effects of environmental and epigenetic factors on the relationships among the three diseases warrant further investigation. Finally, advanced techniques, such as single-cell analysis and spatial omics, hold the potential to uncover the precise mechanisms of genetic signals within specific cell and tissue types, ultimately facilitating the development of personalized therapeutic strategies.

## Methods

### GWAS datasets

We obtained summary statistics for T2D from the Diabetes Meta-Analysis of Trans-Ethnic association studies (DIAMANTE) Consortium, comprising 180,834 cases and 1,159,055 controls ^23^. Summary statistics for AAA were derived from a multi-ancestry meta-GWAS conducted by the AAAGen consortium, including 39,221 cases and 1,086,107 controls ^25^. Summary statistics for CAD were derived from a meta-GWAS involving 210,842 cases and 1,167,328 controls, including data from the UK Biobank, Biobank Japan, and the CARDIoGRAMplusC4D consortium ^24^. GWAS Summary statistics for 15 cardiometabolic traits were derived from the UK Biobank (https://www.ukbiobank.ac.uk/) and numerous large consortia, six immune cell traits from the Blood Cell Consortium (BCX) ^54^, and six immune diseases from FinnGen (https://www.finngen.fi/en) and the GWAS catalog (https://www.ebi.ac.uk/gwas). Detailed information of these GWAS studies is provided in Supplementary Table 1.

### Power calculation for the GWAS

The discovery power of GWAS studies on T2D, CAD and AAA were assessed by GAS Power Calculator (https://csg.sph.umich.edu/abecasis/gas_power_calculator/). Under different minor allele frequency values, we computed the odds ratios (OR) required in each GWAS to achieve a statistical power greater than 0.5, modeling with the numbers of cases and controls reported in each study and disease prevalence rates derived from literature: 0.009 for AAA ^55^, 0.061 for T2D ^56^, and 0.036 for CAD ^57^.

### Genome-wide genetic correlation

Genetic correlations between traits were estimated by linkage disequilibrium score regression (LDSC) ^58^, which quantified separate contributions of polygenic effects by examining the relationship between LD scores and summary statistics of GWAS SNVs. The correlation estimates range from −1 to 1, indicating perfect negative to perfect positive genetic correlation. For this analysis, we utilized pre-computed LD scores obtained from ∼1.2 million common SNVs in the well-imputed HapMap3 European ancestry panel. Bonferroni correction was applied to adjust for multiple testing.

### Local genetic correlation

Local genetic correlations by genomic regions were computed using the Local Analysis of [co]Variant Association (LAVA) ^59^ framework. We downloaded 2,495 semi-independent LD blocks, each approximately 1 Mb in size, from the LAVA repository (https://github.com/cadeleeuw/lava-partitioning) to calculate local genetic heritability and covariance. FDR correction was used to account for multiple comparisons.

Considering that some loci were pleiotropic, we evaluated conditional genetic relationships between phenotypes by employing partial correlation and multivariable regression within LAVA ^60^. Partial correlation computes genetic correlation (r_g_) between two phenotypes when conditioning on other phenotypes, reflecting shared genetic effects attributable to a third phenotype. Separately, multivariable regression models the genetic contribution of multiple predictor phenotypes to an outcome phenotype, enabling us to determine the extent to which genetic components of the predictors explain the genetic signals of the outcome. We used both methods to characterize genetic correlations of some chromosomal segments.

### Cross-trait meta-analysis of GWAS

To identify pleiotropic loci shared between diseases, we performed cross-trait meta-analysis of GWAS summary statistics using Multi-Trait Analysis of GWAS (MTAG) ^61^, and conducted cross-phenotype association analysis (CPASSOC) ^62^ across traits as a sensitivity analysis. MTAG incorporates LDSC to account for sample overlap among GWASs of multiple correlated traits. Its key assumption is that all SNVs share the same variance-covariance matrix of effect sizes across traits. As initially described ^61^, MTAG consistently provides effect estimates with a lower genome-wide mean squared error compared with single-trait GWAS estimates. Furthermore, MTAG-derived association statistics enhance statistical power and minimize FDR inflation for traits with high correlation.

For the CPASSOC analysis, we used Shet’s test. Independent SNVs were identified via PLINK clumping with parameters: --clump-p1 5 × 10^−8^ --clump-p2 1 × 10^−5^ ---clump-r2 0.1 --clump-kb 1000. Pleiotropic SNVs were defined meeting the following criteria: *P* < 5 × 10^−8^ in the meta-analysis (i.e., *P_MTAG_* & *P_CPASSOC_*) and *P* < 1 × 10^-5^ in both GWAS studies.

ANNOVAR was used for functional annotation of the variants identified by MTAG and CPASSOC. The shared SNVs were visualized in a circular dendrogram using the R package ggraph.

### SNV colocalization analysis

Between-trait colocalization was conducted using the R package coloc ^63^ to test whether the associations between two diseases were driven by the SNVs in LD. We extracted the sentinel SNVs and SNVs in ±1 Mb flanking regions for each disease (T2D, CAD or AAA), along with other disease in the same genomic windows. Default priors were used, setting p1 = 1 × 10^−4^, p2 = 1 × 10^−4^, and p12 = 1 × 10^−5^. The colocalization yields posterior probabilities corresponding to one of five hypotheses: PPH0 (no association with either trait); PPH1 (association with gene/protein expression, but not disease risk); PPH2 (association with disease risk, but not gene/protein expression); PPH3 (association with disease risk and gene/protein expression, driven by distinct causal variants); and PPH4 (association with disease risk and gene/protein expression, driven by a shared causal variants) ^63^. To boost the discovery power, we considered genes with PPH3 + PPH4 ≥ 0.8, irrespective of whether the causal variants were shared or distinct.

Multi-trait colocalization was performed by Hypothesis Prioritization for multi-trait Colocalization (HyPrColoc), using the R package HyPrColoc. This Bayesian approach identifies share genetic associations across traits using GWAS summary statistics ^64^. For each *cis*-eQTL sentinel SNV, we extracted them and SNVs in ±1 Mb flanking regions from disease-relevant tissues in GTEx v.8, alongside GWAS summary statistics from corresponding genomic windows. A posterior probability (PP) of ≥ 0.8 was considered strong evidence of colocalization.

### Phenome-wide association study (PheWAS)

Phenome-wide association studies were performed using PheWeb ^65^ (https://pheweb.org/UKB-TOPMed/), which enabled identification of associations between SNVs and a range of phenotypes. We utilized GWAS summary statistics of 786 phenotypes from the UK Biobank, ensuring each GWAS included at least 500 cases.

### Gene-based association analysis and directionality assessment

Identification of the disease-associated GWAS genes was performed using TWAS ^66^, SMR ^67^, MAGMA ^68^, and GCTA-fastBAT ^69^. Each tool was run on GWAS datasets to derive candidate genes (FDR < 0.01), followed by integration using the criterion: (GCTA ⋂ MAGMA) ⋃ (SMR ⋃ TWAS).

Transcriptome-wide association study (TWAS) identifies tissue-specific gene-trait associations by integrating GWAS data with eQTL information ^70^. We conducted TWAS using the FUSION software ^66^, leveraging 43 post-mortem tissue expression profiles in GTEx with pre-computed models ^71^.

Summary-data-based Mendelian Randomization (SMR) integrates GWAS and eQTL datasets to identify gene expressions associated with traits due to pleiotropy or causality ^67^. SMR implements the HEIDI-outlier test to distinguish pleiotropy from linkage. We implemented SMR using *cis*-eQTL summary data from eQTLGen ^72^ (whole blood) and GTEx V8 (nine relevant tissues: artery aorta, adipose subcutaneous, artery coronary, artery tibial, heart atrial appendage, heart left ventricle, kidney cortex, liver, and whole blood). Disease-associated genes were defined as those with *P*_SMR_ passing FDR thresholds and *P*_HEIDI_ >0.05.

We ran MAGMA and GCTA-fastBAT with default parameters, using the European ancestry panel of the 1000 Genomes Project (Phase 3) as the LD reference.

Three methods were used to identify directions of association between genes and diseases: TWAS, SMR, and a weighted average effect formula ^73^:

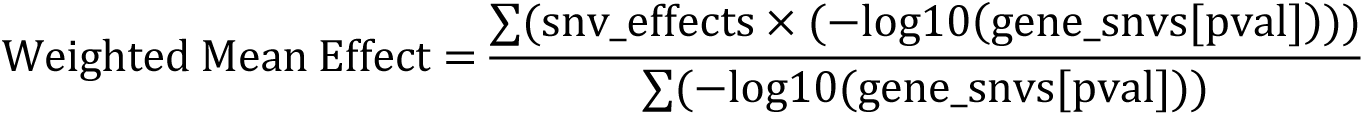

where gene_snvs[pval] represents the *P* value for each SNV of the gene, and snv_effects denotes the SNV estimates.

### Gene expression levels in tissues and cells

mRNA expression levels for genes of interest across 37 tissues were analyzed using GTEx gene expression profiles, as integrated in the Human Protein Atlas (https://www.proteinatlas.org/). Expression levels of these genes were further analyzed across various cell types within tissues.

### GO and KEGG enrichment analysis

Gene Ontology (GO) and Kyoto Encyclopedia of Genes and Genomes (KEGG) pathway enrichment analyses were performed using the “clusterProfiler” package ^74^. The Benjamini–Hochberg procedure was applied to account for multiple testing, with significant pathways defined by FDR < 0.05.

### Differential gene expression analysis

Transcriptomic data were obtained from the Gene Expression Omnibus (GEO) database. For AAA, GSE57691 dataset was used, which contained gene expressions in the aortic tissues from 49 AAA patients and 10 healthy controls. For T2D, GSE38642 and GSE23343 datasets were used. The GSE38642 dataset contained gene expressions in the pancreatic tissue of 9 T2D patients and 54 healthy controls, while the GSE23343 dataset contained gene expressions in the liver tissues of 10 T2D patients and 7 healthy controls. Normalization and differential expression analyses were conducted using the limma R package.

### GTEx tissue-specific expression analysis (TSEA)

We conducted tissue-specific expression analysis (TSEA) ^75^ on the shared genes against all measured genes in GTEx, which contained RNA-Seq data from 1,839 samples across 45 tissues derived from 189 post-mortem individuals. Hypergeometric tests were used to determine whether tissue-specific genes were enriched among the genes shared by two diseases. We used Benjamini–Hochberg correction to account for multiple testing, using the threshold FDR < 0.05.

### Cell type-specific enrichment analysis (CSEA)

To identify the cell types in which the shared genes were enriched for expression, CELLECT ^76^ was used to integrate single-cell RNA sequencing (scRNA-seq) data with cross-trait GWAS summary statistics obtained by MTAG. The scRNA-seq data were obtained from the Human Protein Atlas database (https://www.proteinatlas.org/about/download), including 81 classical cell types from 31 datasets and 45 immune cell types from the Monaco ^77^ and Schmiedel ^78^ datasets. Expression specificity was calculated for each gene within given cell types. We used the established MAGMA model in CELLECT to quantify the associations between GWAS signals and cell type expression specificity. An FDR-corrected *P* value of 0.05 was set as the significance threshold.

We also performed cell type enrichment through WebCSEA ^79^ and Enrichr ^80^, using shared genes as inputs. WebCSEA queries gene sets against tissue-cell-type expression signatures derived from 11 single-cell gene expression datasets. Specifically, Dai et al ^79^ used a t-statistics-based method “deTS” to derive tissue-cell-type signature genes and performed Fisher’s exact test to assess whether the gene set of interest was overrepresented among all cell type-specific genes. For Enrichr (https://maayanlab.cloud/Enrichr/), a web-server tool that contains various datasets, we used the datasets of Azimuth, CellMarker, Descartes, Human Gene Atlas, PanglaoDB and Tabula Sapiens, with CellMarker serving as the primary source to associate shared genes with specific cell types (FDR < 0.05). Results were validated against at least one independent dataset to ensure robustness and biological relevance (*P* value < 0.05).

### Between-trait Mendelian randomization (MR) analysis

Two-sample MR analyses were generally used in our analyses, using the R package TwoSampleMR V.0.5.7. Inverse variance weighting (IVW) was the primary method, with weighted mode, weighted-median and MR Egger regression as sensitivity analyses. For instrument selection, SNVs meeting the significance threshold *P* < 5 x 10^-8^ were selected, with LD clumping performed to identify independent SNVs (r^2^ < 0.001 within 1 Mb) using the 1000 Genomes Project Phase 3 (EUR) as the reference panel. Cochran’s Q test was used to detect heterogeneity of causal estimates, and MR-Egger intercept test was applied to detect potential directional pleiotropy.

For causal inference between metabolic traits and T2D/CAD/AAA, GWAS SNVs for each trait and disease were used as inputs.

For causal inference between T2D and AAA, the instruments for T2D were selected from three input sources: a large GWAS dataset from the Diabetes Meta-Analysis of Trans-Ethnic association studies (DIAMANTE) Consortium involving 180,834 T2D patients and 1,159,055 controls ^23^, a small GWAS dataset involving 34,840 T2D patients and 114,981 controls ^31^, and a set of fine-mapped T2D SNVs, which were initially discovered from 74,124 T2D cases and 824,006 controls and used for MR study of T2D and AAA ^30^.

For causal inference between T2D mechanistic clusters and CAD/AAA, eight T2D mechanistic clusters defined by Suzuki et al ^35^ were obtained. These clusters include beta cell +PI (91 SNVs), beta cell -PI (89 SNVs), residual glycemic (389 SNVs), body fat (273 SNVs), metabolic syndrome (166 SNVs), obesity (233 SNVs), lipodystrophy (45 SNVs), liver and lipid metabolism (3 SNVs). Genetic instruments were then selected within each cluster.

### Disease-disease matching by pathway pairing

Pathway-based disease-disease matching was computed as previously described ^29^. Pairing between the top 20 pathology pathways and other pathology pathways was defined as tier-1, while pairing between the top 20 pathways from one group and the 21^st^ – 50^th^ pathways from the other group constituted tier-2. Pathology pathways were derived by combining GWAS genes with disease genes from databases (MalaCards, DisGeNET, GeneCard, OpenTargets) for pathway enrichment.

### Software packages used

Publicly available software was used to perform the analyses. Analysis software include:

LDSC (https://github.com/bulik/ldsc);

LAVA (https://github.com/josefin-werme/lava);

MTAG (https://github.com/JonJala/mtag);

CPASSOC (;http://hal.case.edu/~xxz10/zhu-web);

PLINK (https://www.cog-genomics.org/plink/1.9) ;

Coloc (;https://github.com/chr1swallace/coloc);

HyPrColoc (https://github.com/cnfoley/hyprcoloc);

PheWAS (https://pheweb.org/UKB-TOPMed/);

TWAS FUSION (http://gusevlab.org/projects/fusion);

MAGMA (https://ctg.cncr.nl/software/magma);

SMR (https://cnsgenomics.com/software/smr/#Overview);

GCTA-fastBAT (https://yanglab.westlake.edu.cn/software/gcta/#fastBAT);

ClusterProfiler (https://github.com/YuLab-SMU/clusterProfiler);

TSEA (http://doughertylab.wustl.edu/tsea);

CELLECT(https://github.com/perslab/CELLECT);

WebCSEA (https://bioinfo.uth.edu/webcsea);

Enrichr (https://github.com/wjawaid/enrichR).

## Data availability

GWAS summary statistics for this study was listed in Supplementary Table 1. The eQTL summary data for eQTLGen and GTEx are available from https://www.eqtlgen.org/cis-eqtls.html and https://gtexportal.org/home/datasets.

## Acknowledgements

S.Z., Y.W. and A.L are Ph.D. students at School of Life Sciences, Fudan University. This study was supported by the research grants awarded to C.P. from National Natural Science Foundation of China (No. 32270626) and Greater Bay Area Institute of Precision Medicine (Guangzhou) (I0005). We thank Drs. Weijia Zhang and Yuxiang Dai from Fudan University, and members of the Laboratory of Intelligent Computing in Biomedicine in the Greater Bay Area Institute of Precision Medicine (Guangzhou) for insightful discussions and suggestions.

## Author contributions

C.P. and S.Z. designed the study. S.Z., Y.W., A.L. and Z.W. performed bioinformatic and statistical analyses. S.Z., Y.W., A.L. and C.P. generated the figures and tables. C.P., T.P.S, J.X. and W.H. provided critical biological insight in interpreting the results. C.P. and S.Z. drafted the manuscript. All authors critically reviewed the manuscript.

## Abbreviation

| Category | Abbreviations | Full name |
| --- | --- | --- |
| Traits | T2D | type 2 diabetes mellitus |
|  | CAD | coronary artery disease |
|  | AAA | abdominal aortic aneurysm |
|  | TAA | thoracic aortic aneurysm |
|  | FI | fasting insulin |
|  | FG | fasting glucose |
|  | 2hGlu | 2-hour glucose |
|  | HbA1c | hemoglobin A1C |
|  | HDLC | high-density lipoprotein cholesterol |
|  | LDL-C | low-density lipoprotein cholesterol |
|  | TG | triglycerides |
|  | TC | total cholesterol |
|  | nonHDLC | non-high-density lipoprotein cholesterol |
|  | SBP | systolic blood pressure |
|  | DBP | diastolic blood pressure |
|  | PP | pulse pressure |
|  | BMI | body mass index |
|  | WHR | waist-hip ratio |
|  | WHRadjbmi | BMI-adjusted waist-hip ratio |
| Gene/Protein | ADAMTS7 | ADAM Metallopeptidase with Thrombospondin Type 1 Motif 7 |
|  | APOC1 | Apolipoprotein C1 |
|  | APOE | Apolipoprotein E |
|  | BSN | Bassoon Presynaptic Cytomatrix Protein |
|  | CDKN2B-AS1 | Cyclin-Dependent Kinase Inhibitor 2B Antisense RNA 1 |
|  | CELSR2 | Cadherin EGF LAG Seven-Pass G-Type Receptor 2 |
|  | CMIP | C-Maf Inducing Protein |
|  | DRD4 | Dopamine Receptor D4 |
|  | HMGCR | 3-Hydroxy-3-Methylglutaryl-CoA Reductase |
|  | IL18BP | Interleukin 18 Binding Protein |
|  | KCNJ11 | Potassium Inwardly Rectifying Channel Subfamily J Member 11 |
|  | LPL | Lipoprotein Lipase |
|  | NUCB2 | Nucleobindin 2 |
|  | PCSK9 | Proprotein Convertase Subtilisin/Kexin Type 9 |
|  | PSRC1 | Proline/Serine-Rich Coiled-Coil 1 |
|  | SLC22A3 | Solute Carrier Family 22 Member 3 |
|  | SMAD3 | SMAD Family Member 3 |
|  | SORT1 | Sortilin 1 |
|  | TANK | TRAF Family Member Associated NFKB Activator |
|  | TCF7L2 | Transcription Factor 7 Like 2 |
|  | TOMM40 | Translocase Of Outer Mitochondrial Membrane 40 |

---

**Supplementary Figure 1.**
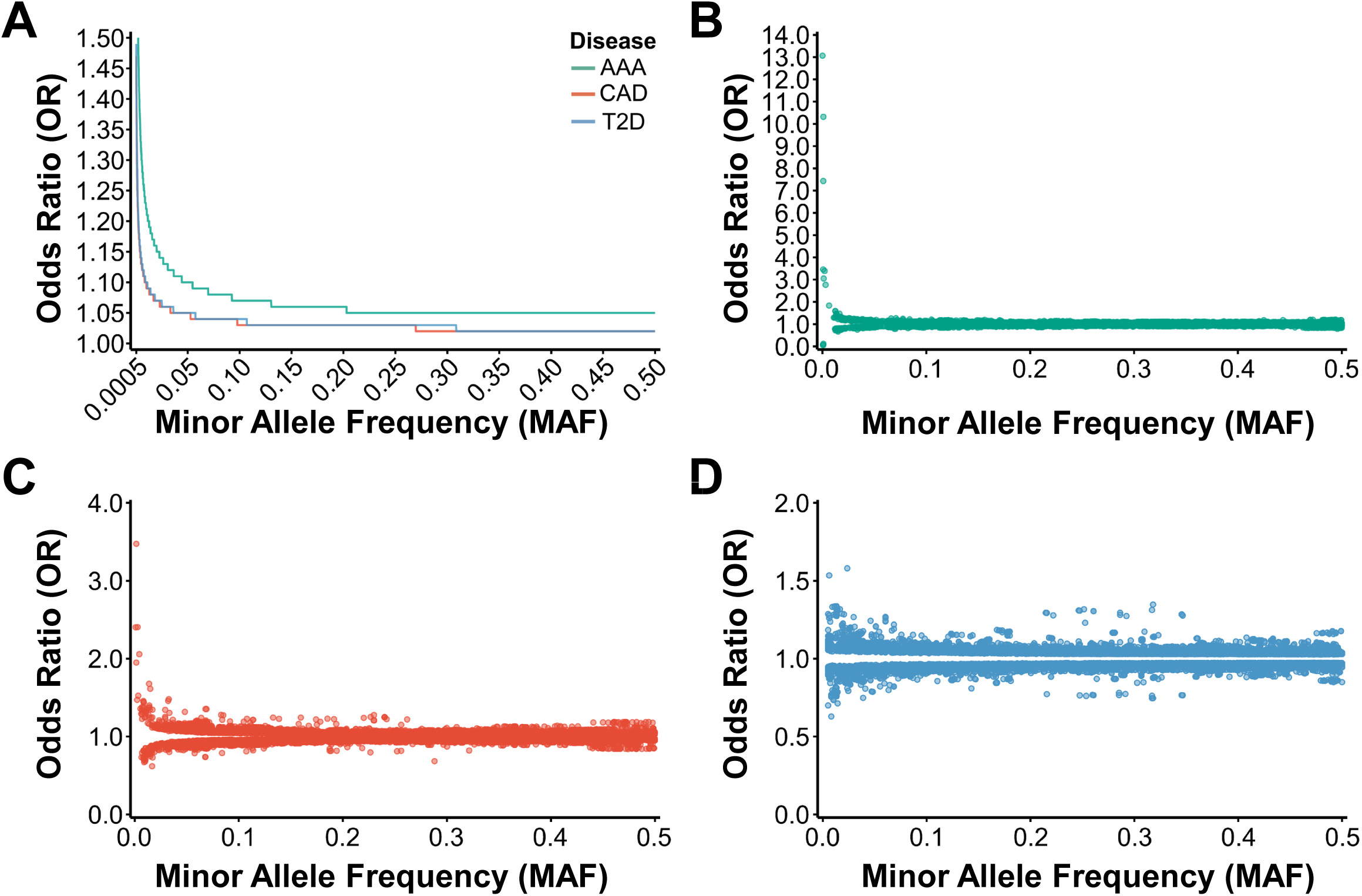
Power assessment for the GWAS datasets. **(A)** Modeling discovery power of 0.5 for the GWAS datasets of T2D (blue line), CAD (red line), and AAA (green line), under different minor allele frequency settings. **(B)** Distribution of SNVs in the AAA GWAS dataset. **(C)** Distribution of SNVs in the CAD GWAS dataset. **(D)** Distribution of SNVs in the T2D GWAS dataset.

**Supplementary Figure 2.**
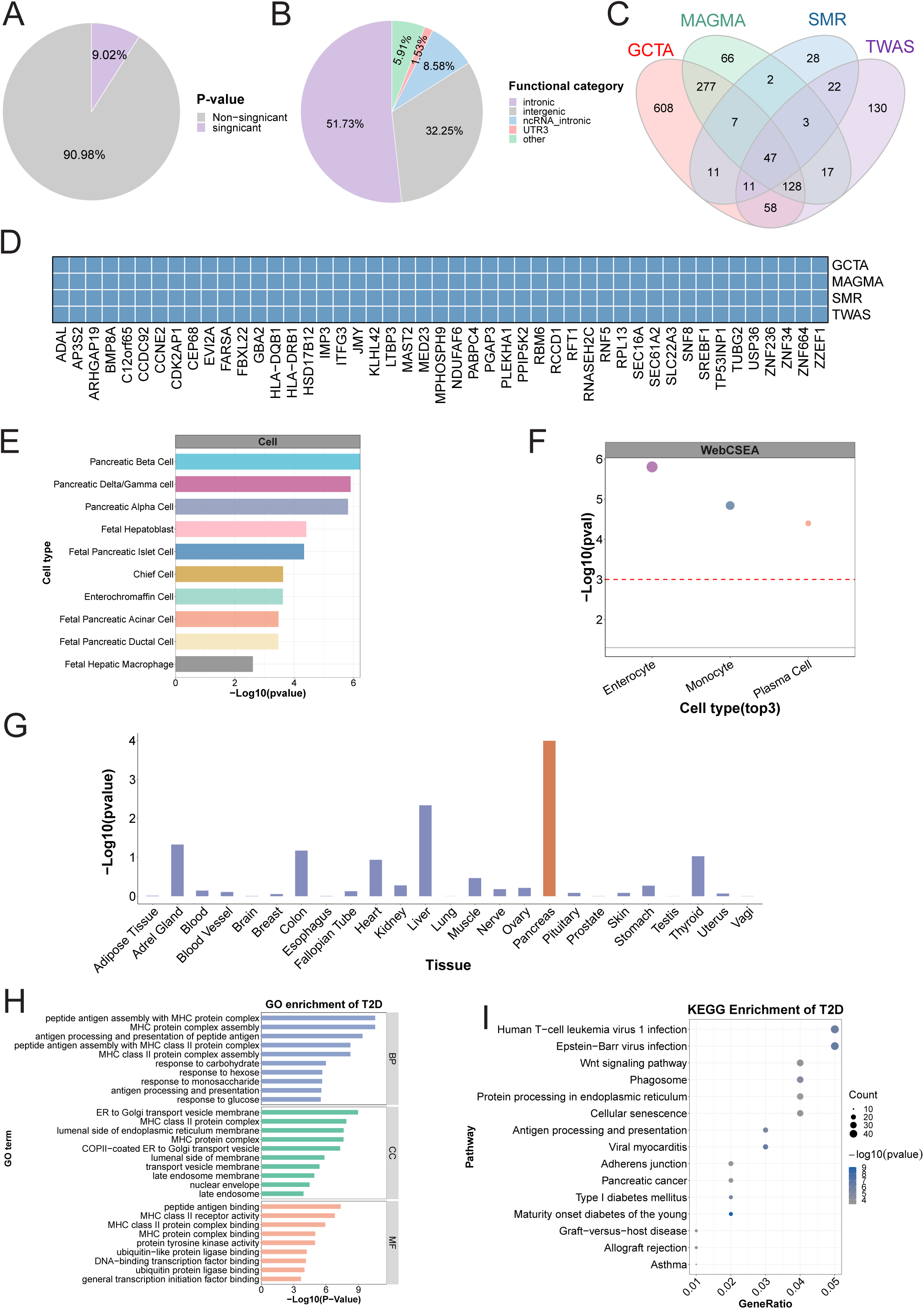
Interpretation of GWAS loci for T2D. **(A)** Distribution of GWAS P values. **(B)** Genic locations of significant SNVs (GWAS P < 5 ×10^-8^). **(C)** The number of T2D-related genes identified by four gene-based analysis methods. **(D)** T2D-related genes identified by all four gene-based association analyses. **(E)** Cell-type enrichment based on SNV heritability localized to cell type-specific enhancers derived from CATLAS, using S-LDSC. **(F)** Cell-type enrichment based on cell type-specific genes defined by various single-cell transcriptome studies, computed via Cell Specific Expression Analysis (CSEA). The top three enriched cell types are shown. **(G)** Tissue enrichment based on tissue-specific genes defined by GTEx, computed via Tissue Specific Expression Analysis (TSEA). Orange represents significantly enriched tissues passing the Bonferroni corrected P value threshold. **(H)** GO enrichment analyses based on the identified T2D-related genes. **(I)** KEGG enrichment analyses based on the identified T2D-related genes.

**Supplementary Figure 3.**
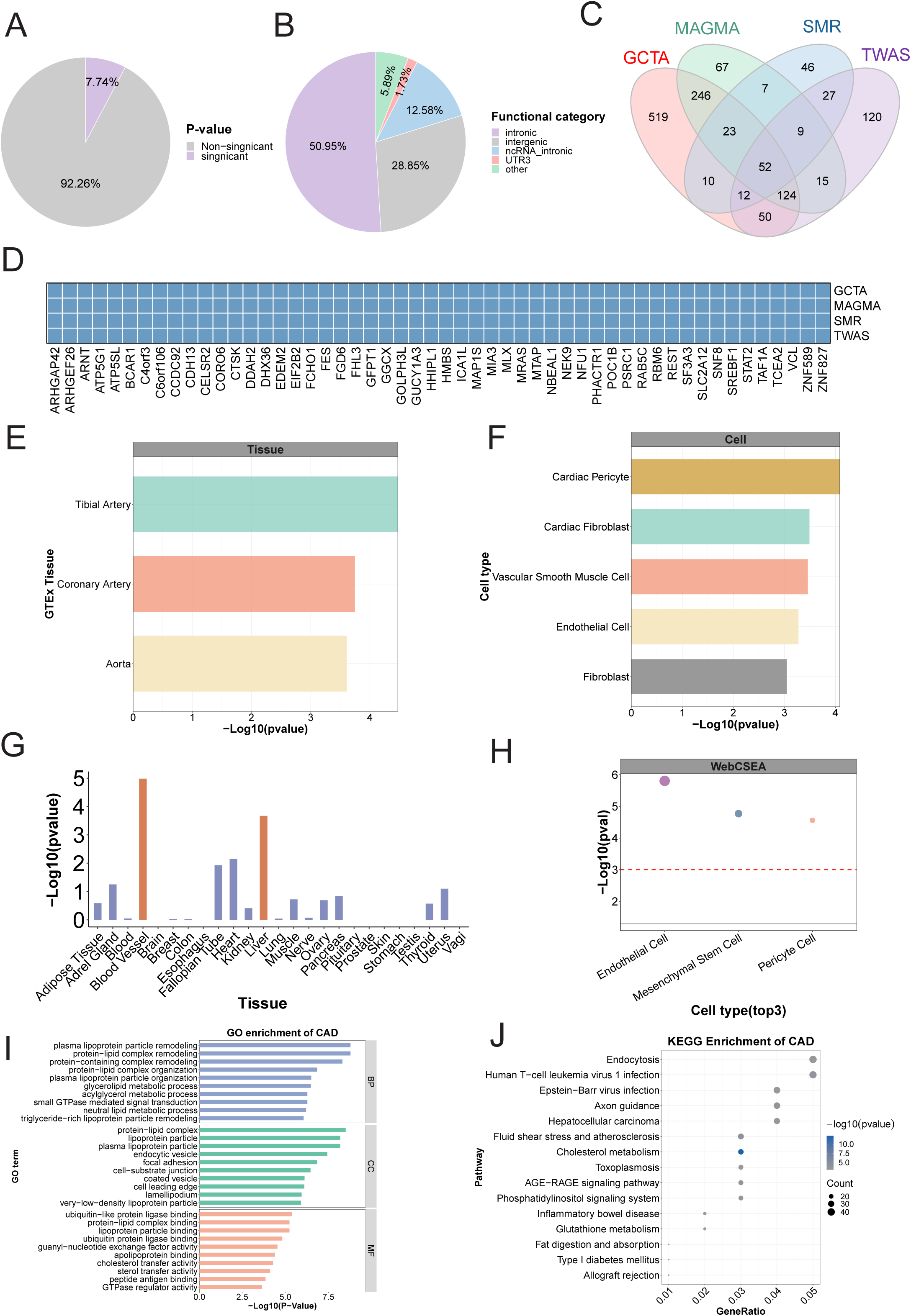
Interpretation of GWAS loci for CAD. **(A)** Distribution of GWAS P values. **(B)** Genic locations of significant SNVs (GWAS P < 5 ×10^-8^). **(C)** The number of CAD-related genes identified by four gene-based analysis methods. **(D)** CAD-related genes identified by all four gene-based association analyses. **(E)** Tissue enrichment based on SNV heritability in tissue-specific genes defined by GTEx, computed by S-LDSC. **(F)** Cell-type enrichment based on SNV heritability localized to cell type-specific enhancers derived from CATLAS, using S-LDSC. **(G)** Tissue enrichment based on tissue-specific genes defined by GTEx, computed via Tissue Specific Expression Analysis (TSEA). Orange represents significantly enriched tissues passing the Bonferroni corrected P value threshold. **(H)** Cell-type enrichment based on cell type-specific genes defined by various single-cell transcriptome studies, computed via Cell Specific Expression Analysis (CSEA). The top three enriched cell types are shown. **(I)** GO enrichment analyses based on the identified CAD-related genes. **(J)** KEGG enrichment analyses based on theidentified CAD-related genes.

**Supplementary Figure 4.**
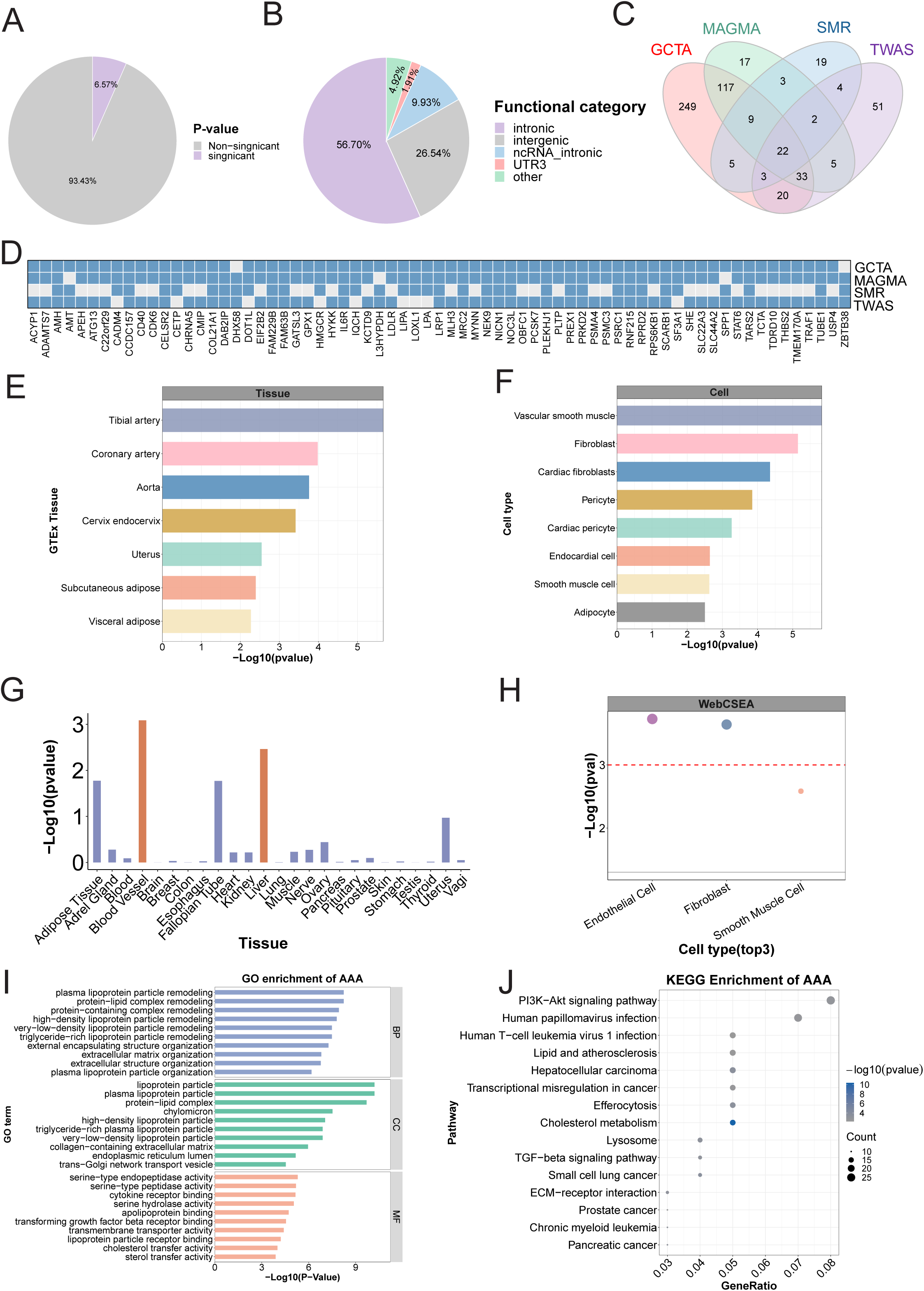
Interpretation of GWAS loci for AAA. **(A)** Distribution of GWAS P values. **(B)** Genic locations of significant SNVs (GWAS P < 5 ×10^-8^). **(C)** The number of AAA-related genes identified by four gene-based analysis methods. **(D)** AAA-related genes identified by more than three of the four gene-based association analyses. **(E)** Tissue enrichment based on SNV heritability in tissue-specific genes defined by GTEx, computed by S-LDSC. **(F)** Cell-type enrichment based on SNV heritability localized to cell type-specific enhancers derived from CATLAS, using S-LDSC. **(G)** Tissue enrichment based on tissue-specific genes defined by GTEx, computed via Tissue Specific Expression Analysis (TSEA). Orange represents significantly enriched tissues passing the Bonferroni-corrected P value threshold. **(H)** Cell-type enrichment based on cell type-specific genes defined by various single-cell transcriptome studies, computed via Cell Specific Expression Analysis (CSEA). The top three enriched cell types are shown. **(I)** GO enrichment analyses based on the identified AAA-related genes. (J) KEGG enrichment analyses based on the identified AAA-related genes.

**Supplementary Figure 5.**
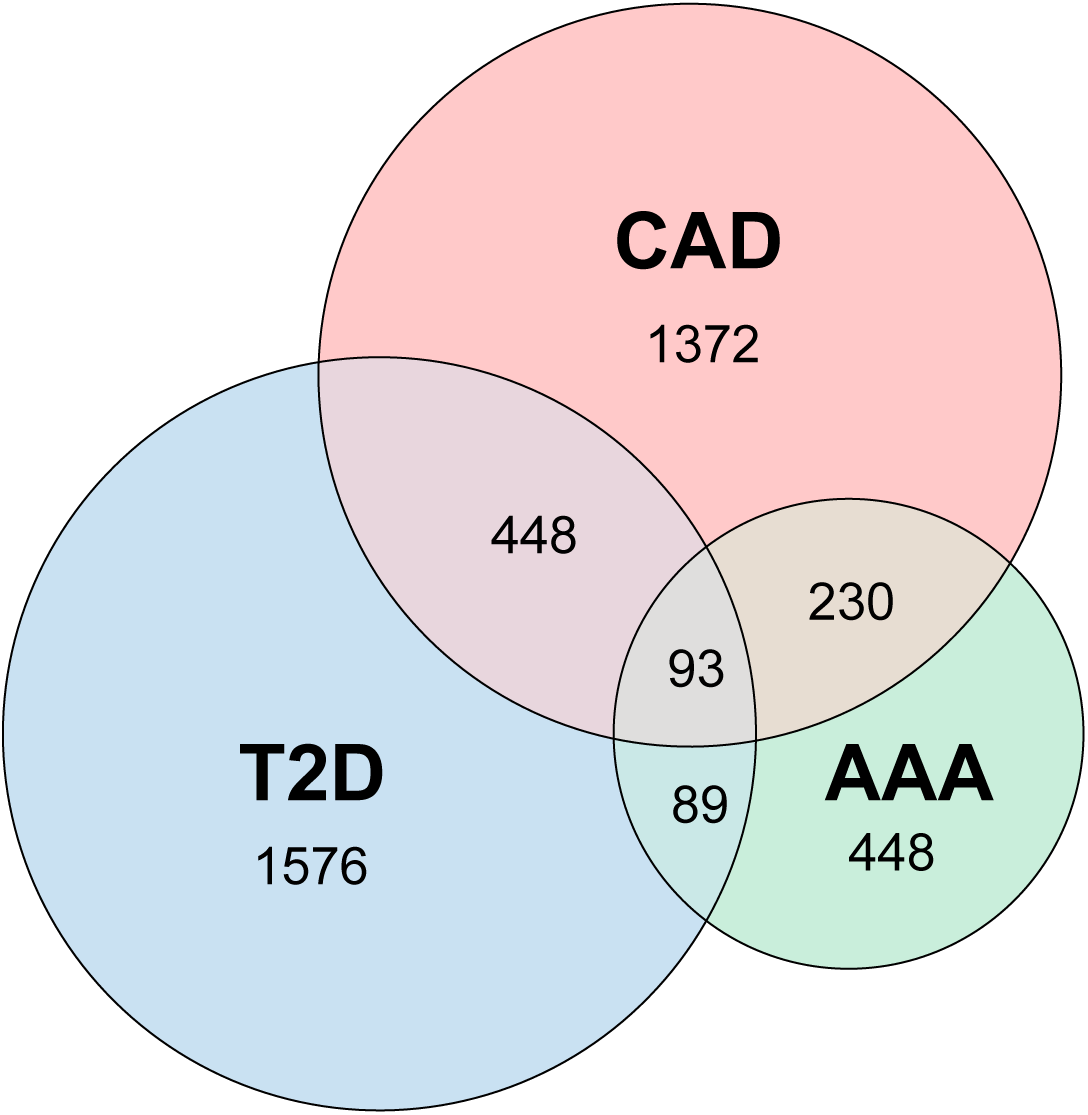
GWAS genes and their overlaps for T2D, CAD and AAA.

**Supplementary Figure 6.**
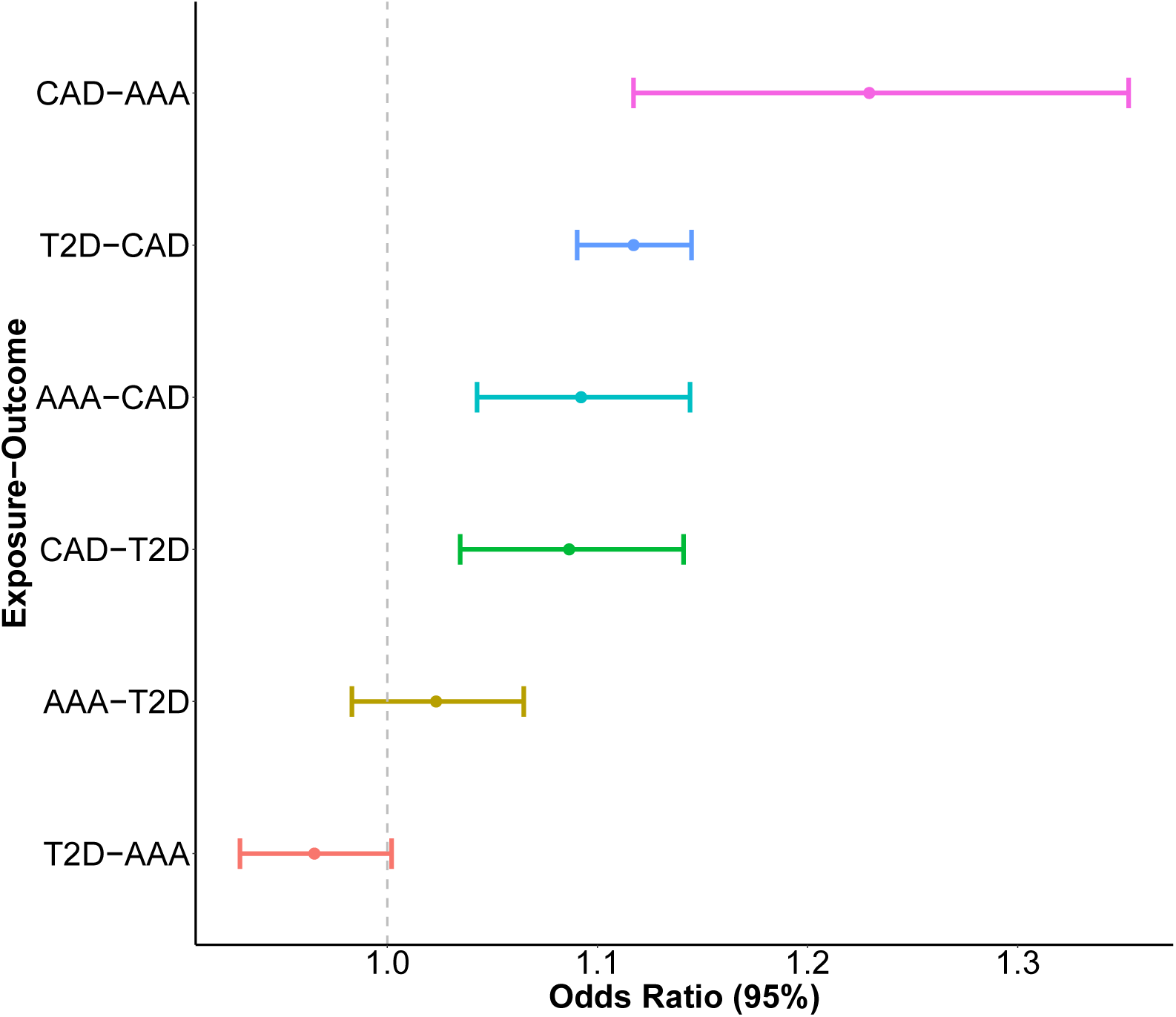
Mendelian randomization to infer causal relationship between the disease pairs. The disease pairs are labeled in the format of exposure – outcome.

**Supplementary Figure 7.**
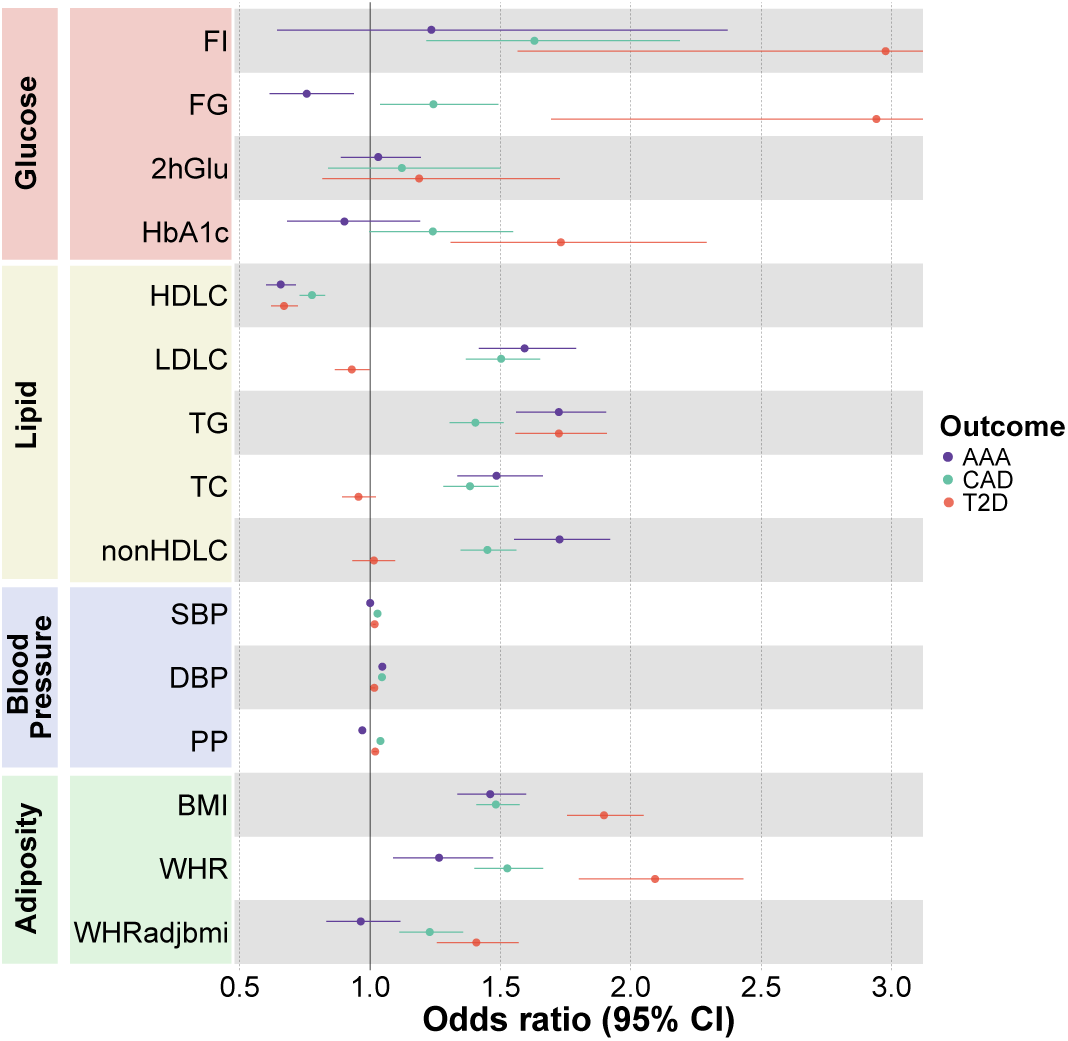
Mendelian randomization to infer causal effects of the metabolic traits on T2D, CAD and AAA.

**Supplementary Figure 8.**
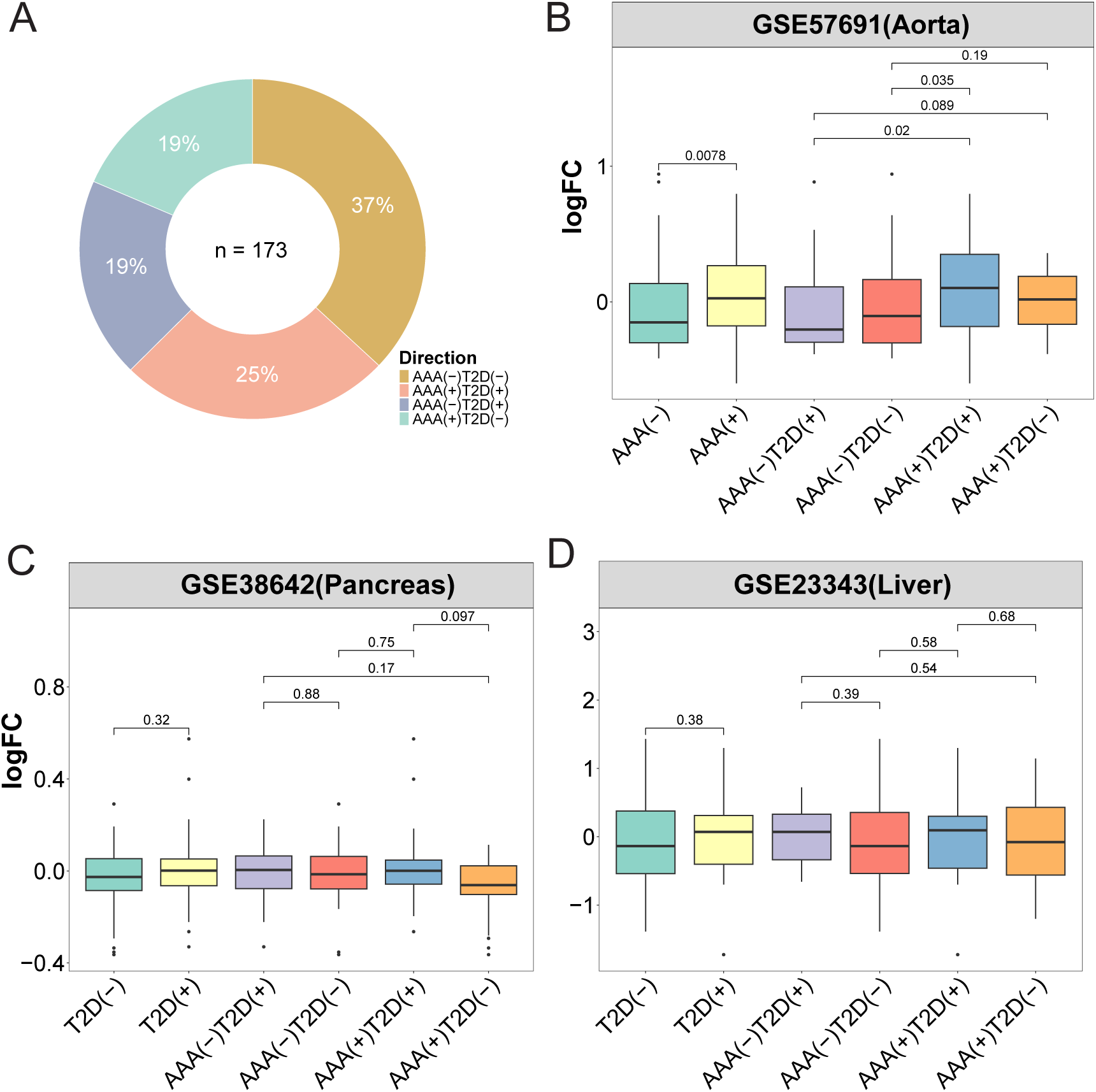
Classification of T2D-AAA shared genes by disease association directions. **(A)** Four groups of genes are identified based on the directions of association with T2D and AAA. Gene expression levels are analyzed in the aorta of the AAA patients **(B),** the pancreas of the T2D patients **(C)**, and the liver of the T2D patients **(D)**. Expression differences between groups are evaluated using the wilcoxon rank-sum test, with corresponding P values indicated above the boxplots.

**Supplementary Figure 9.**
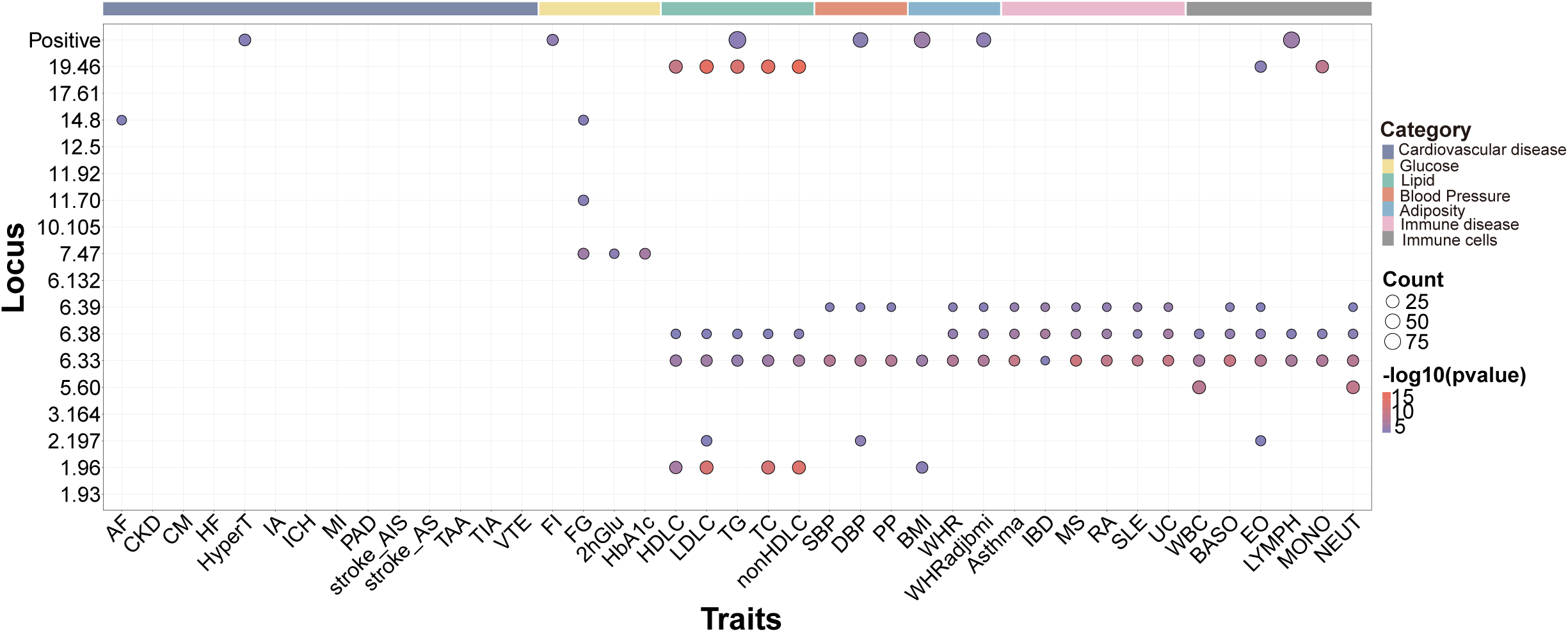
Enrichment of GWAS genes in the 17 chromosomal segments where T2D and AAA display negative genetic correlations. The GWAS traits contain 7 categories, including 14 cardiovascular diseases, 15 metabolic traits (4 glucose traits, 5 blood lipid traits, 3 blood pressure traits, and 3 adiposity traits), 6 immune diseases, and 6 immune cell types (cell abundance levels).

**Supplementary Figure 10.**
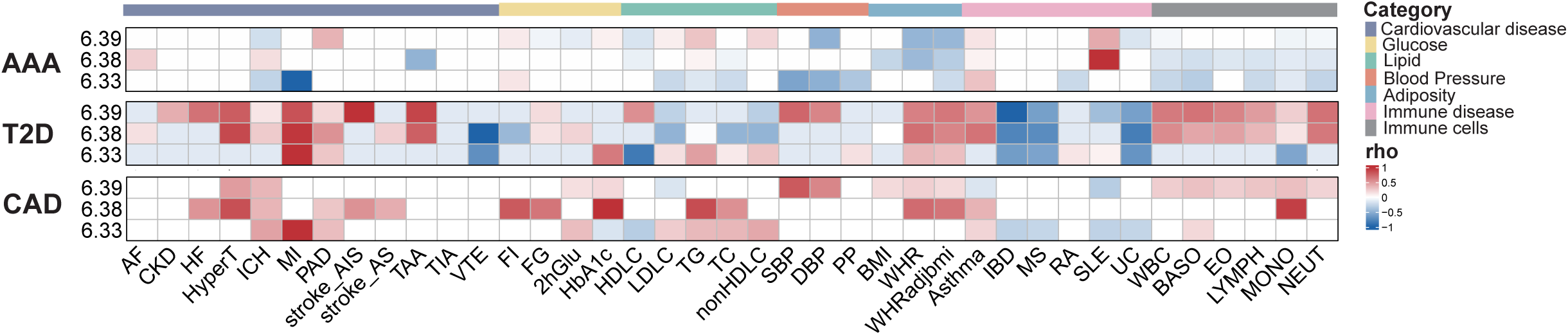
T2D, AAA and CAD display distinct correlation profiles with cardiometabolic and immune traits across three HLA regions. rho: genetic correlation.

**Supplementary Figure 11.**
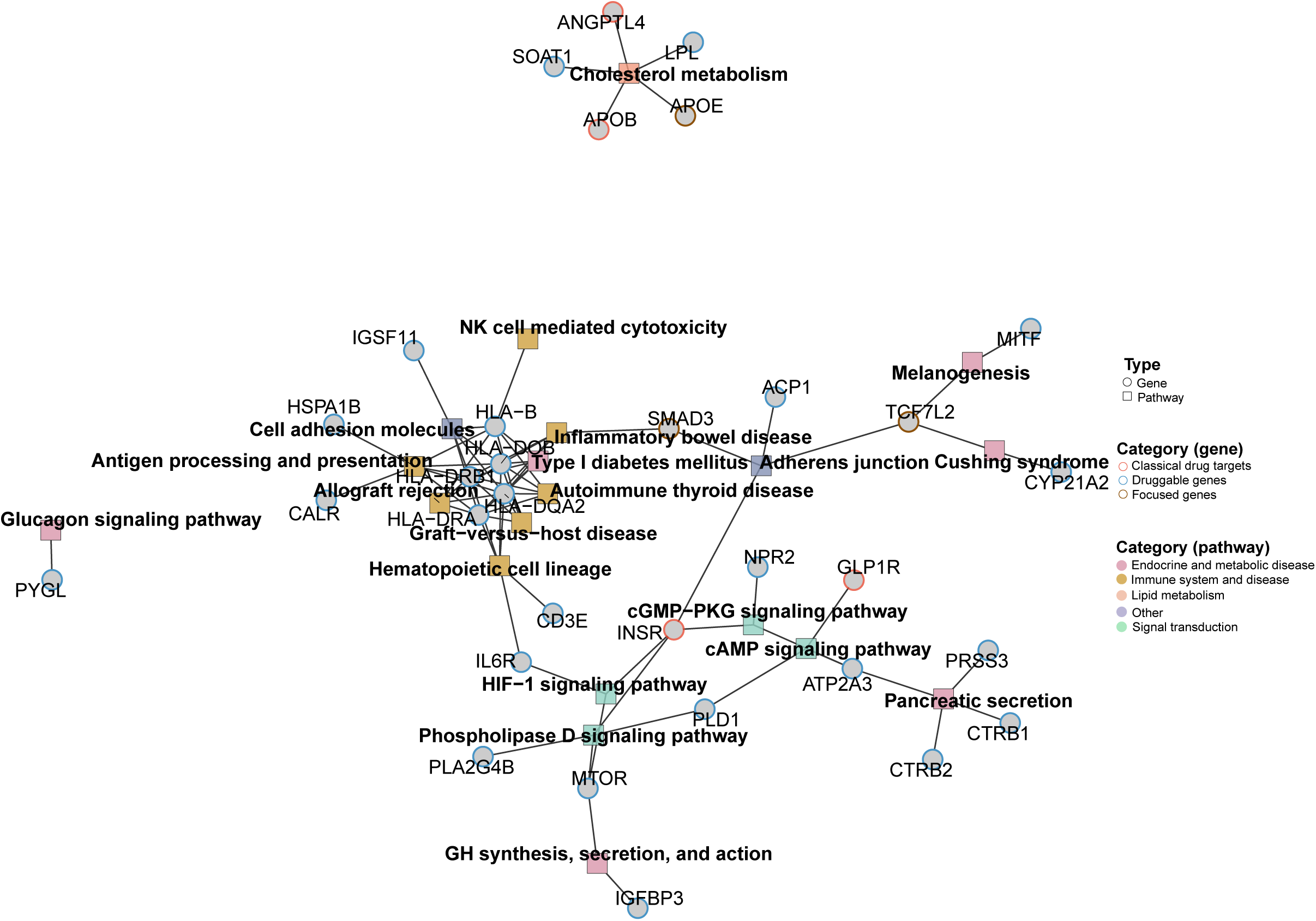
Pathway connections for important genes exhibiting opposite associations with T2D and AAA. Genes are presented as dots, with outer circles denoting their classification: targets of classical drugs identified in drug-gene databases (red), derived from scanning druggable genes and proteins (blue), or genes reported from literature to play important roles in disease pathogenesis (brown). Pathways are presented as squares.

**Supplementary Figure 12.**
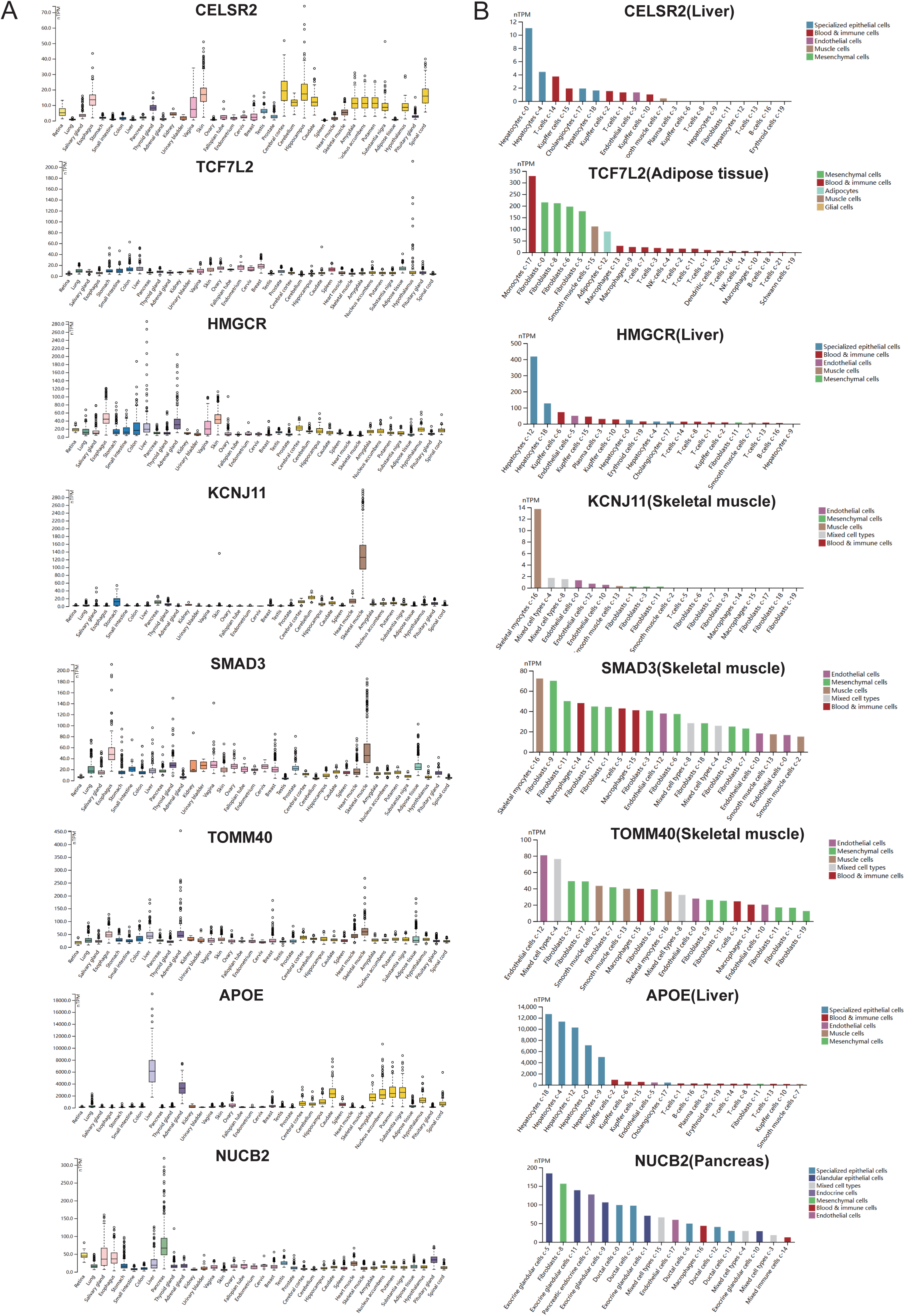
Gene expression across GTEx tissues (A) and cells (B). X axis: tissue types or cell types. Y axis: normalized expression values in TPM from RNA sequencing. For panel B (cell types), the source tissues are annotated next to the gene namess in the figure titles.

**Supplementary Figure 13.**
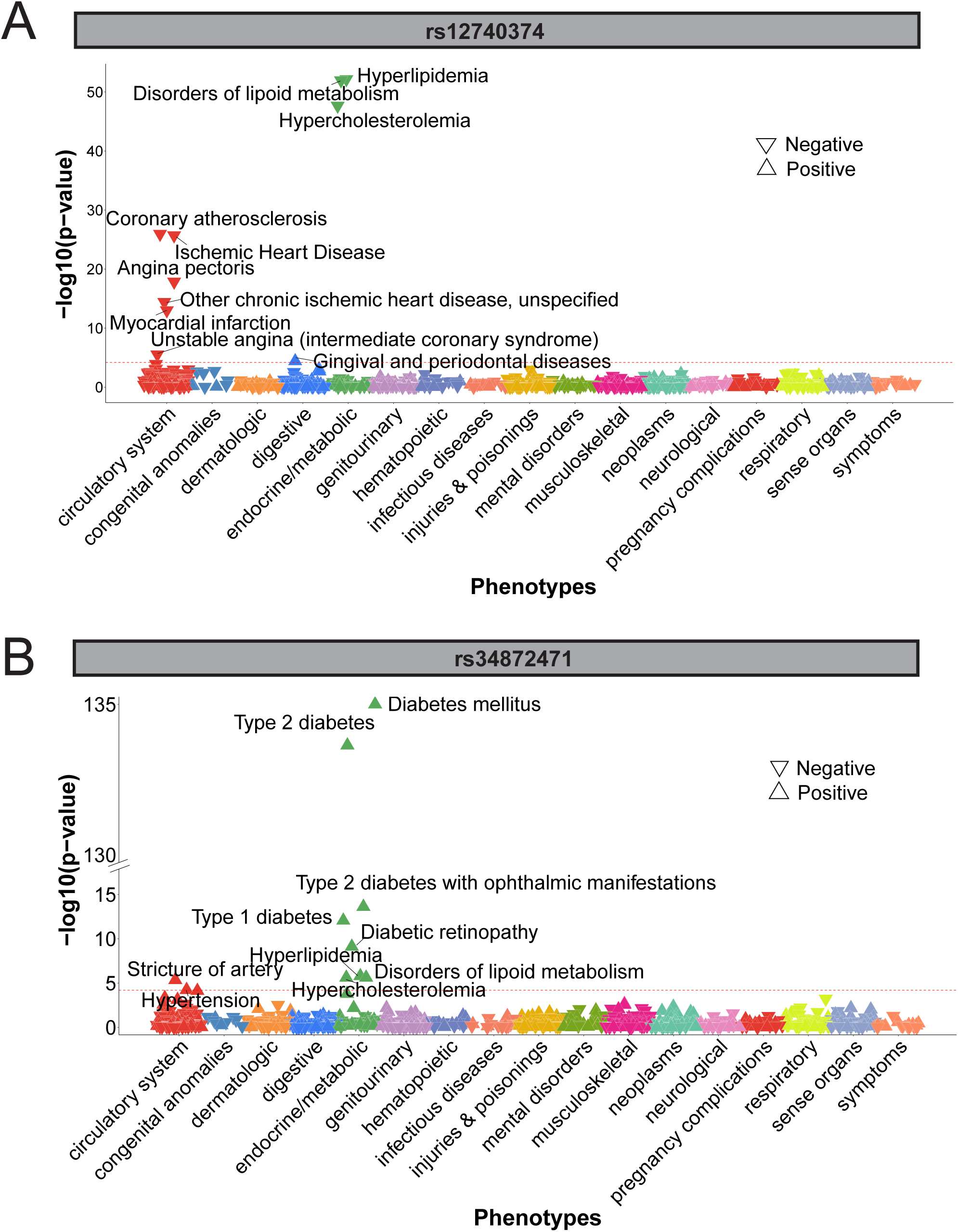
Phenome-wide association studies on two causal SNVs that display opposite associations with T2D and AAA. (A) 12740374, and (B) rs34872471. The association directions, negative or positive, are indicated by triangles. The red dashed line marks the Bonferroni-corrected P value threshold (P value = 6.35 x 10^-5^). Significantly associated phenotypes are denoted.

